# Factors associated with severe acute respiratory syndrome-related coronavirus 2 infection in unvaccinated children and young adults

**DOI:** 10.1101/2023.08.13.23294036

**Authors:** Sarah L. Silverberg, Hennady P. Shulha, Brynn McMillan, Guanyuhui He, Amy Lee, Ana Citlali Márquez, Sofia R. Bartlett, Vivek Gill, Bahaa Abu-Raya, Julie A. Bettinger, Adriana Cabrera, Daniel Coombs, Soren Gantt, David M. Goldfarb, Laura Sauvé, Mel Krajden, Muhammad Morshed, Inna Sekirov, Agatha N. Jassem, Manish Sadarangani

## Abstract

**BACKGROUND:** Pediatric COVID-19 cases are often mild or asymptomatic, which has complicated estimations of disease burden using existing testing practices. We aimed to determine the age-specific population seropositivity and risk factors of SARS-CoV-2 seropositivity among children and young adults during the pandemic in British Columbia (BC).

**METHODS:** We conducted two cross-sectional serosurveys: phase 1 enrolled children and adults <25 years between November 2020-May 2021 and phase 2 enrolled children <10 years between June 2021-May 2022 in BC. Participants completed electronic surveys and self-collected finger-prick dried blood spot (DBS) samples. Samples were tested for immunoglobulin G antibodies against ancestral spike protein (S). Descriptive statistics from survey data were reported and two multivariable analyses were conducted to evaluate factors associated with seropositivity.

**RESULTS:** A total of 2864 participants were enrolled, of which 95/2167 (4.4%) participants were S-seropositive in phase 1 across all ages, and 61/697 (8.8%) unvaccinated children aged under ten years were S-seropositive in phase 2. Overall, South Asian participants had a higher seropositivity than other ethnicities (13.5% vs. 5.2%). Of 156 seropositive participants in both phases, 120 had no prior positive SARS-CoV-2 test. Young infants and young adults had the highest reported seropositivity rates (7.0% and 7.2% respectively vs. 3.0-5.6% across other age groups).

**CONCLUSION:** SARS-CoV-2 seropositivity among unvaccinated children and young adults was low in May 2022, and South Asians were disproportionately infected. This work demonstrates the need for improved diagnostics and reporting strategies that account for age-specific differences in pandemic dynamics and acceptability of testing mechanisms.

## BACKGROUND

Children represented 15-20% of confirmed cases of severe acute respiratory syndrome coronavirus 2 (SARS-CoV-2) infections and less than 0.5% of deaths during the coronavirus disease 2019 (COVID-19) pandemic. Children and young adults accounted for approximately 1,000 intensive care unit admissions in Canada between January 2020 and March 2023.[1, 2] Pediatric estimates of SARS-CoV-2 infection burden have been underestimated; possible reasons include frequent asymptomatic or paucity of symptomatic disease, aversion of children to invasive sampling and lack of non-invasive sampling approaches (e.g. saliva) for the youngest children. There has been limited population-based research on asymptomatic pediatric SARS-CoV-2 incidence or overall prevalence of infection, particularly longitudinally.[3–6] Population-based studies are needed to understand the role of children in COVID-19 and identify highest risk populations, as this has been useful for setting public health guidance around schooling and congregate activities, and may be valuable for prevention strategies for COVID-19 and other respiratory viruses. Notably, BC had one of the earliest provincial returns to in-person schooling in Canada, with children returning to the classroom in June 2020, with virtual learning opportunities still available and planned public health measures including learning groups implemented for the in-person experience.[7] Post-secondary institutions returned in-person in fall, 2021. Seropositivity in BC since then therefore reflects children and youth with the opportunity to participate in congregate activities.

Understanding risk factors for and rates of disease spread among unvaccinated children and youth is representative of many children and youth worldwide. In this analysis from an ongoing COVID-19 seroprevalence and immunity study, we sought to provide additional data on the prevalence of SARS-CoV-2 infection in unvaccinated children and young adults (up to age 25 years) in British Columbia (BC), Canada. In BC, COVID-19 vaccines were generally available for adults aged over 18 years from April 2021, adolescents aged 12-17 years from May 2021, children aged 5-11 years in November 2021, and for younger children ages 6 months to 5 years from August 2022. We estimated age-specific SARS-CoV-2 seropositivity based on presence of anti-spike (S) protein Immunoglobulin G (IgG) antibodies in dried blood spot (DBS) samples, and identified sociodemographic factors associated with seropositivity.

## MATERIALS AND METHODS

### Study population

We conducted two cross-sectional serosurveys of children and young adults less than 25 years of age in BC. BC has a population of approximately 5.3 million individuals, including approximately 1.2 million in the study’s target age group.[8]

All BC residents aged under 25 years were eligible for study participation. There were no specific exclusion criteria other than age and self-reported COVID-19 vaccination status. Study activities were divided into two phases: data were collected in phase 1 from November 15, 2020, to May 1, 2021, and in phase 2 between May 30, 2021, and May 20, 2022. For analytical purposes, all participants from phase 1 and only children aged under 10 years from phase 2 were included to maintain a consistent focus on SARS-CoV-2 infections in unvaccinated individuals; phase 2 analytic restrictions were implemented due to provincial COVID-19 vaccination programs. We additionally divided the study into three time periods for analysis, reflecting the dominant variants of concern circulating, with the first period reflecting ancestral and pre-Delta variants (April 2020-June 2021), the second reflecting the emergence of the Delta variant (June 2021-December 2021), and the third reflecting the emergence of Omicron variant (December 2021 through end of study). Associated public health interventions are described in Supplementary figure 1.

### Data collection

Once consented (and assented as appropriate), data were collected via an electronic survey using REDCap electronic data capture tools hosted at BC Children’s Hospital Research Institute. Participants or caregivers completed the survey directly. Data collected included: socio-demographic data including any known pre-existing medical condition; travel outside British Columbia and/or Canada; potential COVID-19 exposures for the participant and household; current and past symptoms of respiratory illness since January 2020; SARS-CoV-2 testing and COVID-19 diagnosis and treatment history since January 2020 (Supplementary figure 2).[9] Testing by either polymerase chain reaction or at-home rapid antigen test were both recorded as part of SARS-CoV-2 testing history. We used the forward sortation code (first three digits of the postal code) to determine geographical distribution or the sample and categorized patient by regional health authority.

For the infection prevention and control behaviours, participants rated the frequency of use of handwashing, physical distancing, and masking on a five-point Likert scale, with 5 representing use every time, and 1 representing never using them.

### Sample collection and analysis

Participants were sent at-home DBS collection kits to enable a single DBS sample collection, using Whatman 903 Protein Saver Card and a contact-activated lancet. Samples were collected by finger-prick, except for infants aged less than six months, where a heel prick was used. All necessary equipment and instructions to collect samples, as well as a pre-paid envelope for sample return, were included. Participants were instructed to return completed DBS cards immediately via regular mail, and once received they were stored at −80°C. DBS samples were processed by punching four 6mm circular punches and eluted overnight in 350ul of elution buffer.[10]

Ten microliters of DBS eluate were diluted in Meso Scale Discovery (MSD) Diluent 100 at 1:500. Assays were conducted at the provincial reference laboratory at the BC Centre for Disease Control (BCCDC) using the MSD V-PLEX COVID-19 Coronavirus Panel 2 (IgG) to measure concentrations of SARS-CoV-2 S protein antibodies using the MSD QuickPlex SQ120 instrument and reported in arbitrary units per milliliter (AU/mL). Positivity anti-S threshold was defined as 75 AU/mL.[10, 11]

### Ethical approval

Ethics approval was obtained from the University of British Columbia Children’s and Women’s Research Ethics Board (H20-01886-A011).

### Statistical analysis

#### Sample Size

Enrollment for each phase was based on five, 5-year age brackets (0-4, 5-9, 10-14, 15-19, 20-24). There was a target sample size of 500 participants per 5-year age group, per phase; each phase was completed when target sample size was reached. This provided over 95% confidence to detect seroprevalence rates of 5% ± 2%. Although the pre-specified sample size for the study was applicable separately to each phase, we also conducted post-hoc analyses of seropositivity rates within phase 2, to determine power corresponding with circulation of different SARS-CoV-2 variants of concern.

#### Data analysis

All phase 1 participants and unvaccinated phase 2 participants aged under ten years were eligible for inclusion in the analysis reported here. In addition, participants without valid anti-S results were excluded from analysis, including those who returned no or an insufficient sample. Categorical variables were compared using Pearson’s chi-square or Fisher’s exact tests as indicated. For the infection prevention and control behaviours, responses were grouped into three: scores 4 or 5 (most or every time), neutral scores of 3, and scores of 1 or 2 (rarely or never).

Two multivariable analyses were conducted to explore demographic factors associated with SARS-CoV-2 seropositivity. Age, sex, ethnicity, health region, pre-existing medical condition, and travel, were included in the models a priori. We analyzed the effect of time in a separate multivariable model. Variables with p-value <0.1 from univariate models were included in the initial multivariable model and an Akaike Information Criterion based backward elimination procedure was applied; any variables with an adjusted p-value of <0.05 were retained in the final model. Adjusted odds ratios and 95% confidence intervals (CI) were calculated to identify factors associated with seropositivity. Multi-variable analyses only included participants with non-missing data for the relevant variables. Seronegative participants from phase 1 were not excluded from participating again for phase 2; these 457 participants were included independently in analysis in each phase.

All analyses were completed using the R software environment (R [Version 4.1.3; The R Project for Statistical Computing, Vienna, Austria]). The STrengthening the Reporting of OBservational studies in Epidemiology (STROBE) reporting guideline for cross-sectional studies was used (Supplementary table 1).

## RESULTS

### Description of study participants

A total of 2864 unvaccinated participants were included in the analysis (Figure 1), of which 1535 (54%) identified as women/girls (Table 1). Most participants with known postal code resided in the more urban ‘Lower Mainland’ area of BC (Vancouver Coastal Health and Fraser Health Authority regions [1681/2182; 77%]) (Table 1; Supplementary figure 3). Only 126/2864 (4.4%) of participants reported not leaving the house for work or school. Participants reported a mean of 3.2 other household members. Only 408/2864 (14.2%) of respondents reported at least one known COVID-19 exposure, with exposures commonly occurring at school, college, or university (144/612, 23.5%), in contact with friends (129/612, 21.1%) or at home (128/612, 20.9%), though 1894/2864 (66.1%) reported one or more illnesses since January 2020 (Table 1).

**Figure 1:**
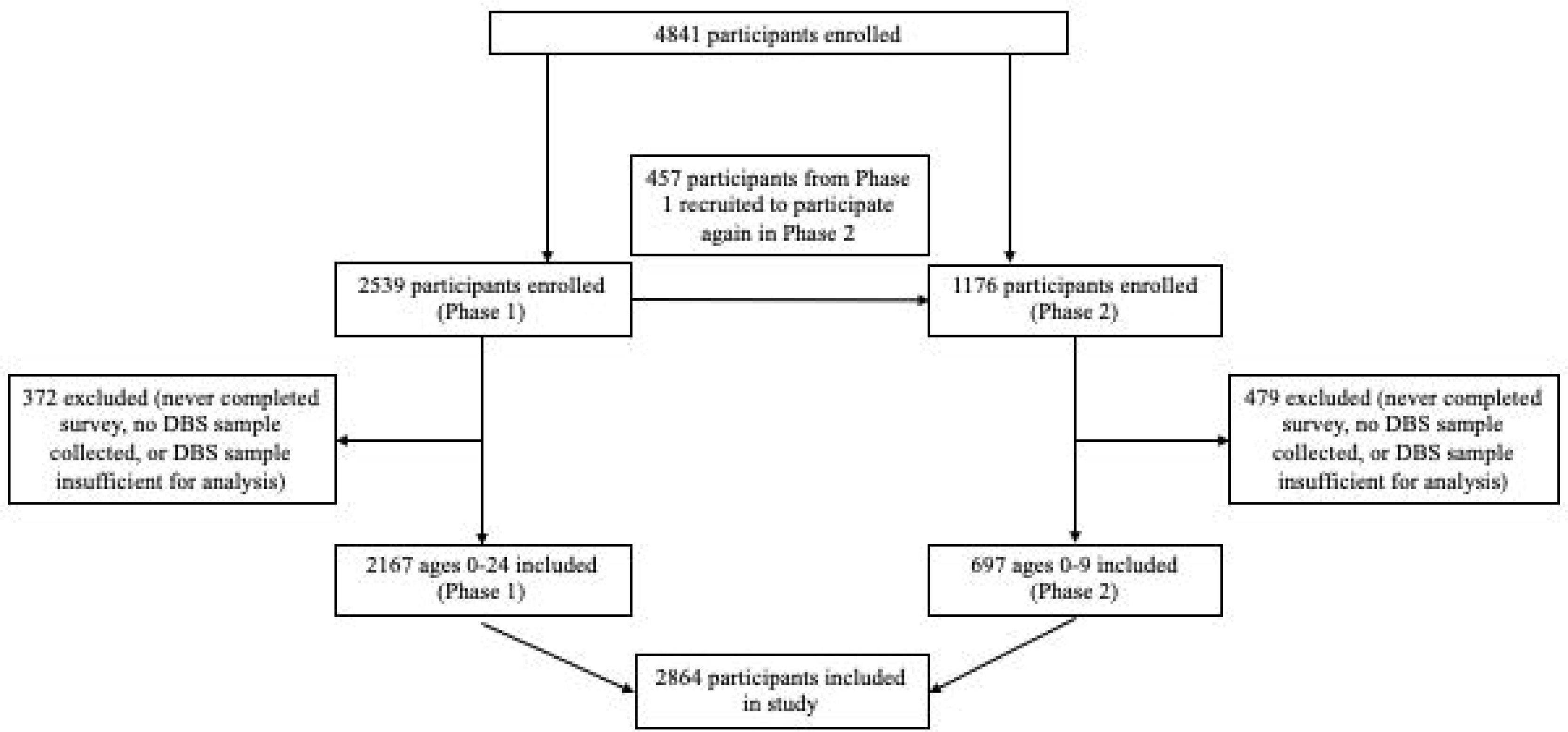
Study Enrollment and Participation. Enrollment of participants across both phases of the study; all participants from phase 1 and only children aged under 10 years from phase 2 were included to maintain a consistent focus on SARS-CoV-2 infections in unvaccinated individuals; phase 2 analytic restrictions were implemented due to provincial COVID-19 vaccination programs.

**Table 1:**
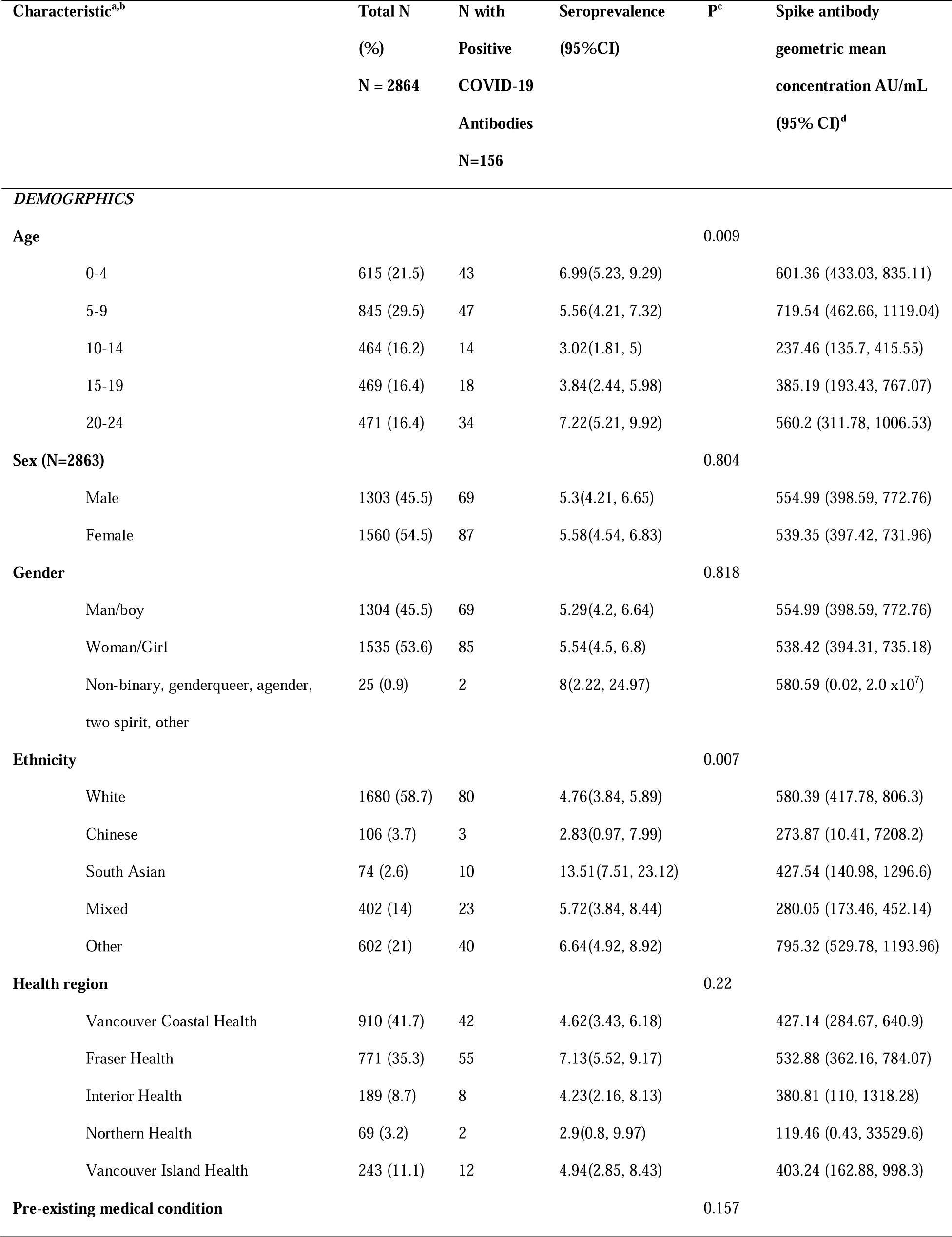
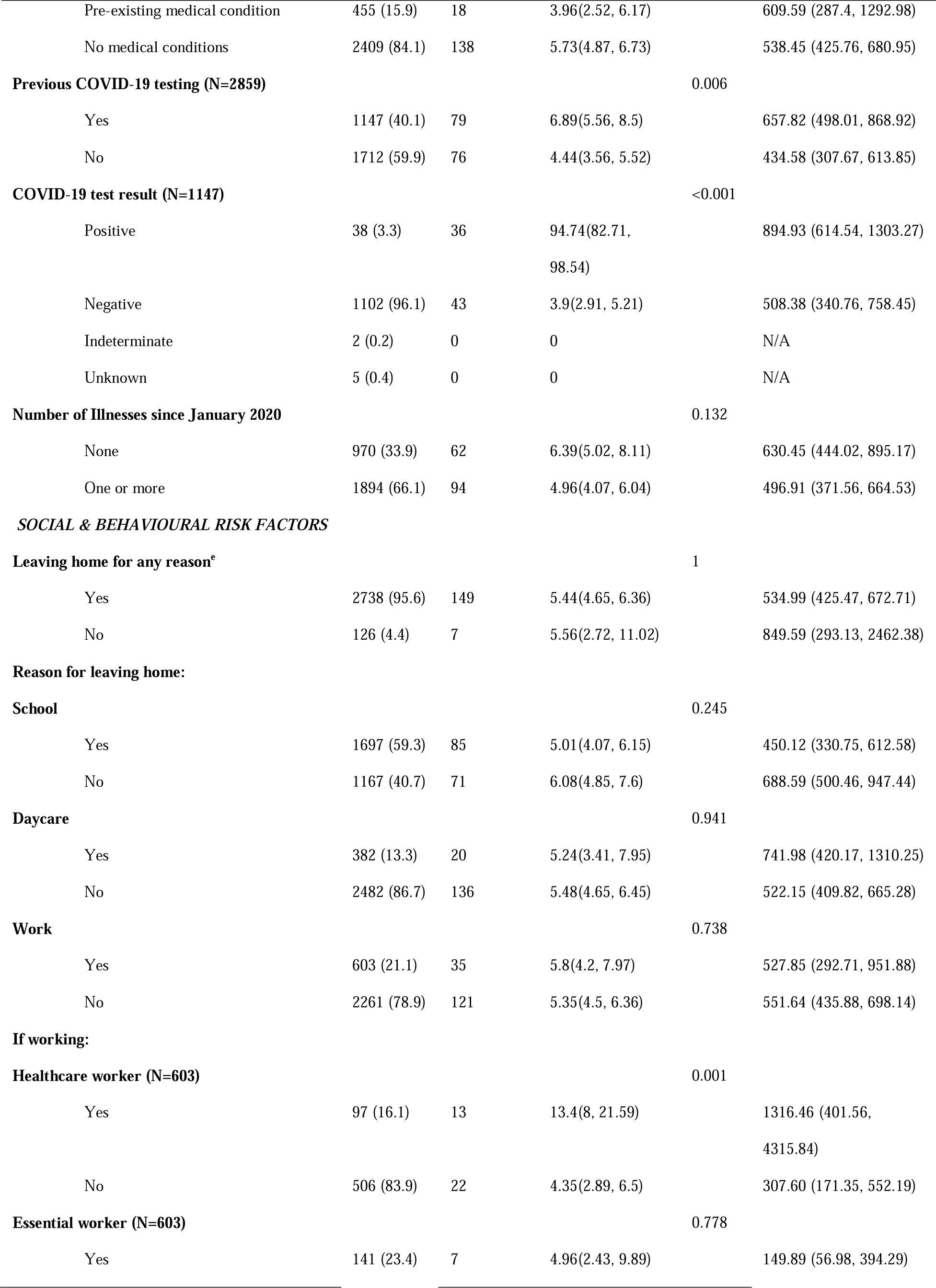
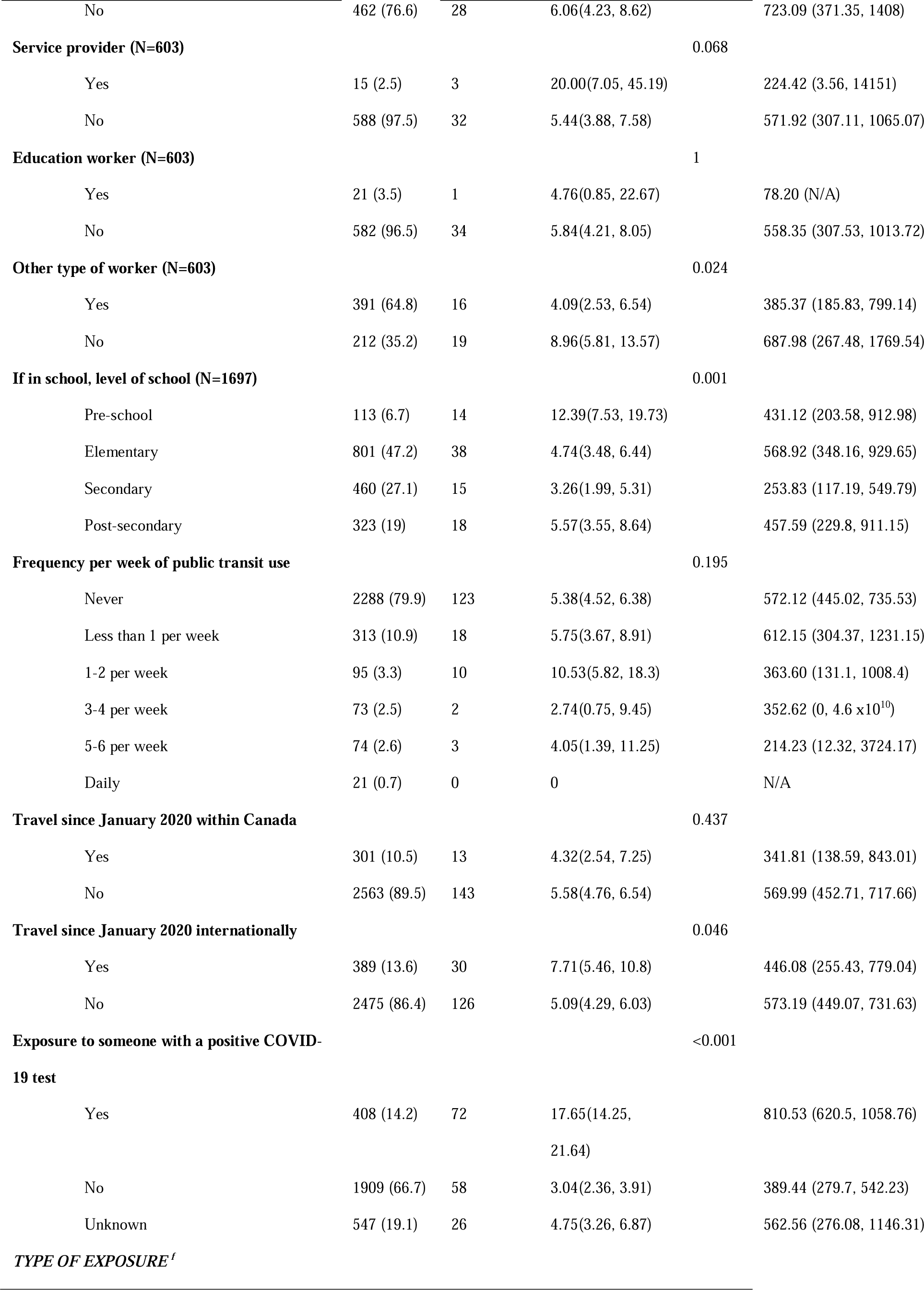
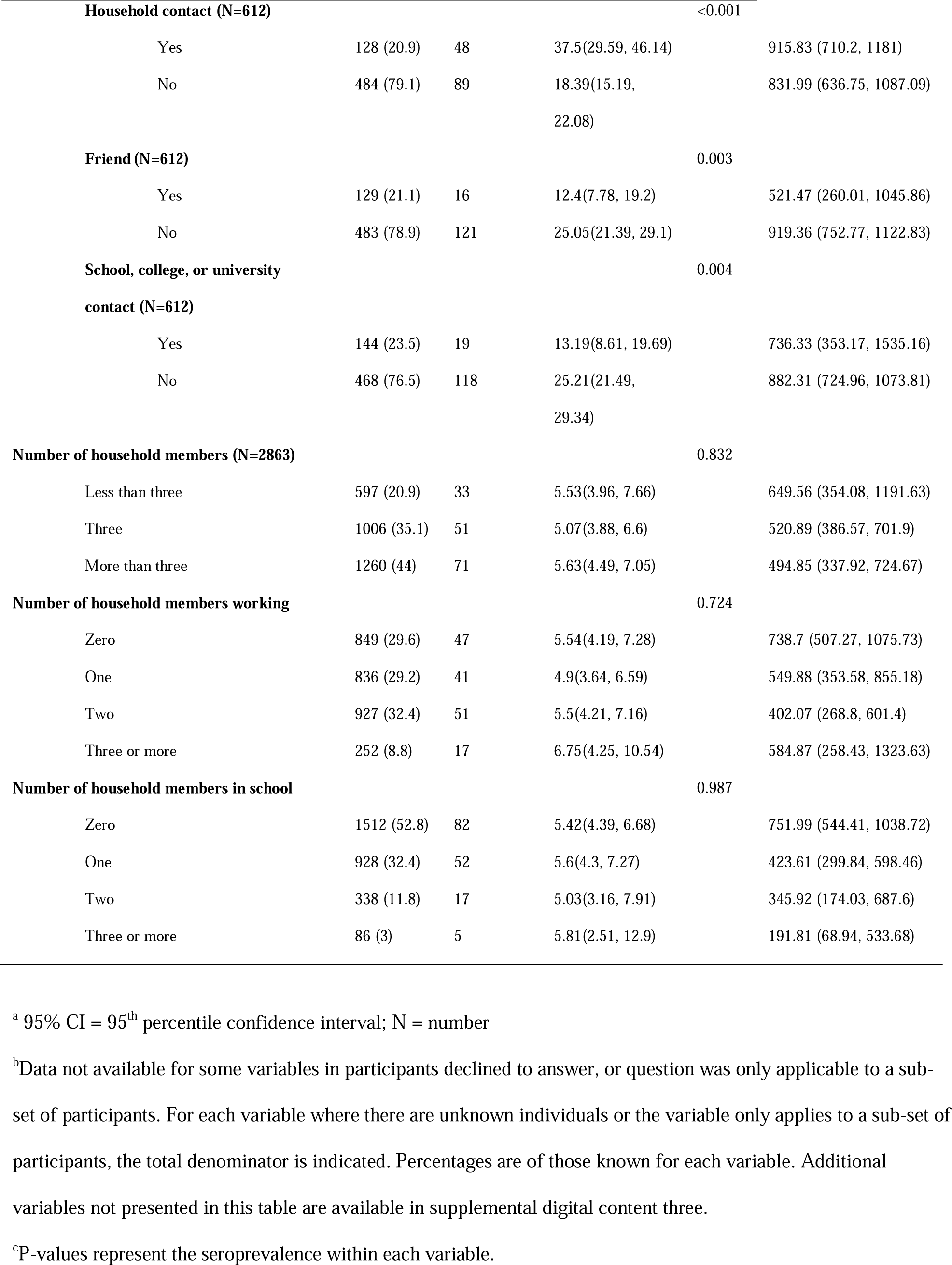
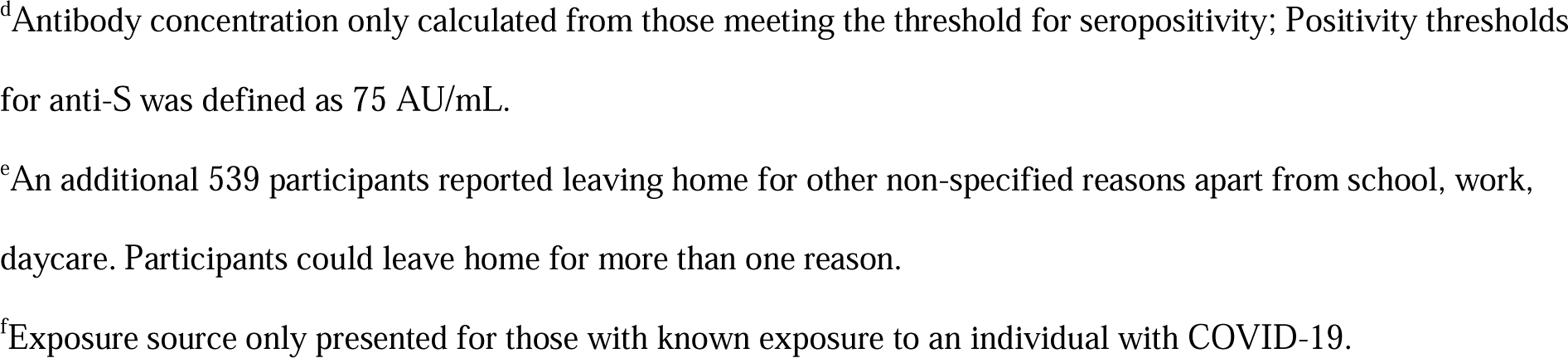
Demographic data.

Participants had a mean score of 4.5/5 for handwashing, 4.0/5 for physical distancing, and 4.2/5 for masking (Supplementary table 2).

### Factors associated with seropositivity

Overall seropositivity was 5.5% (95% CI: 4.7, 6.3). Overall seropositivity for those aged under 10 years in phase 1 was 3.8% (95% CI: 2.7, 5.4), and was 8.8% (95% CI: 6.9, 11.1) during phase 2.

Seropositivity was below 10% for children under ten years of age through December 2021, and was 21.9% (95% CI: 14.0, 32.7) for children aged 0–4 years and 60% (95% CI: 31.3, 83.2) for those aged 5-9 years sampled between January-May 2022 (Figure 2, Table 2).

**Figure 2:**
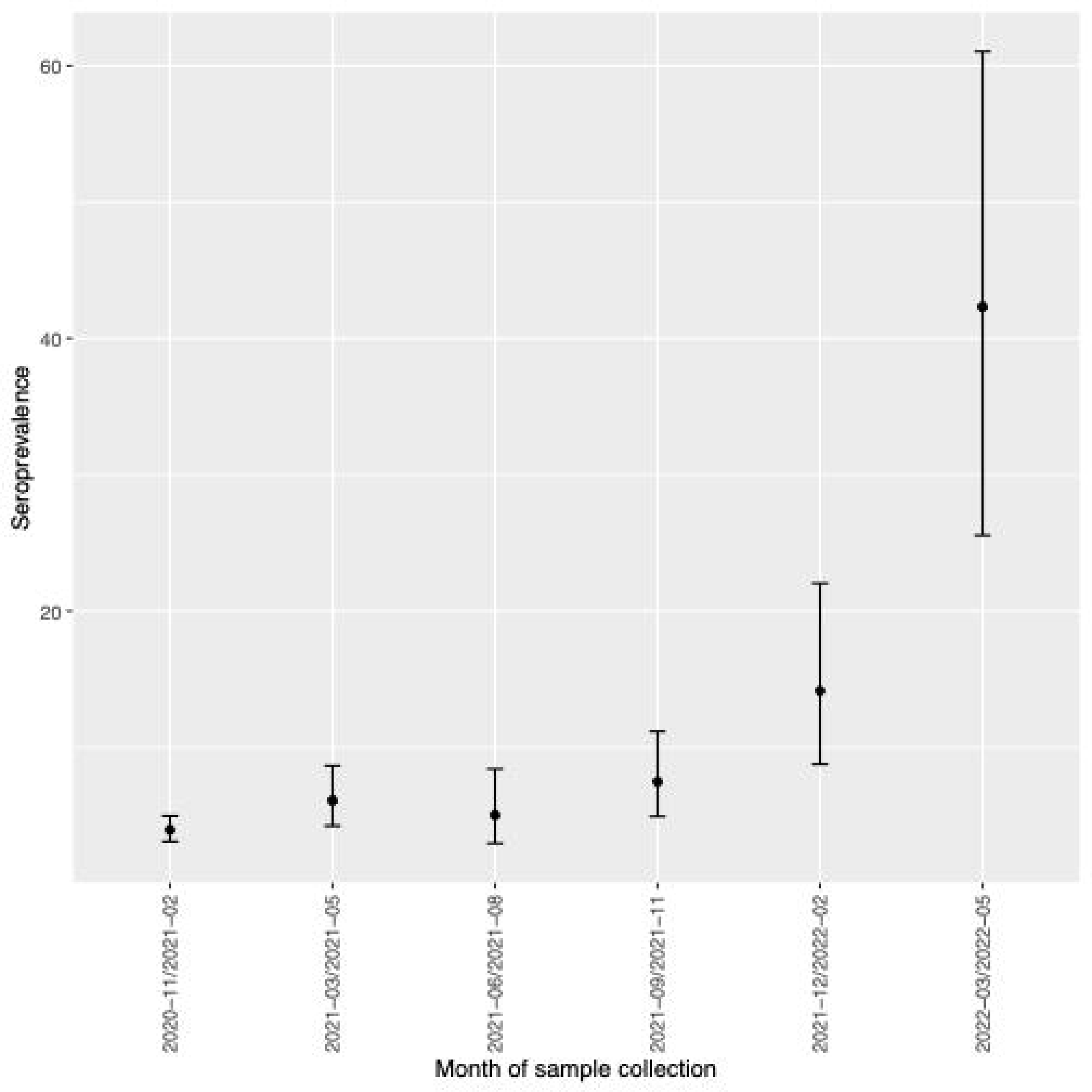
SARS-CoV-2 Anti-S IgG Seroprevalence by date of DBS sample amongst children and young adults in British Columbia between November 2020 and May 2022. Overall SARS-CoV-2 seroprevalence for unvaccinated children and youth in British Columbia, Canada between November 2020 and May 2022. Children and youth under 25 years of age were included from November 2020-May 2021; data from June 2021 and onward representative of children aged under ten years. Aggregated seroprevalence calculated out of all samples collected in the calendar month.

**Table 2:**
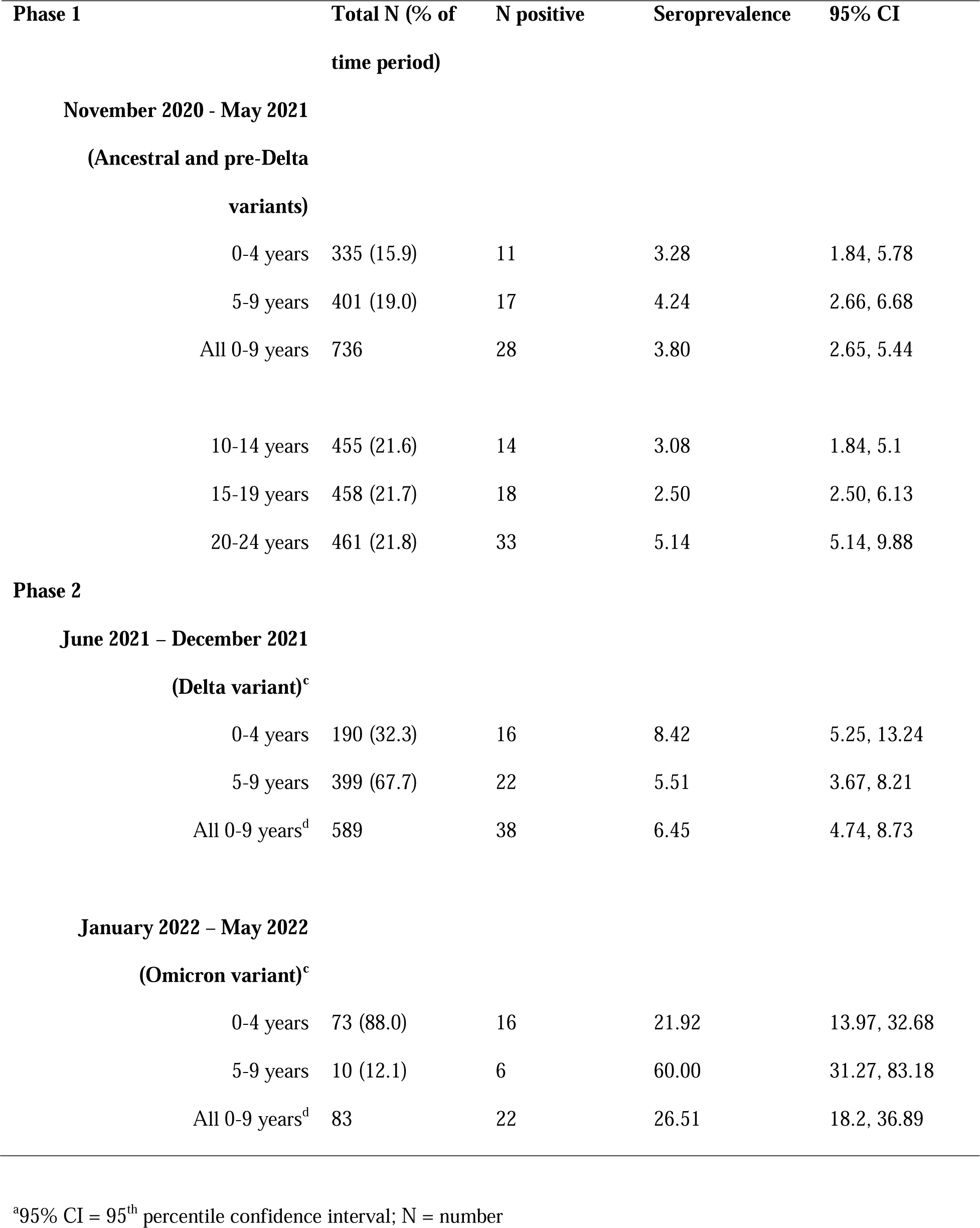
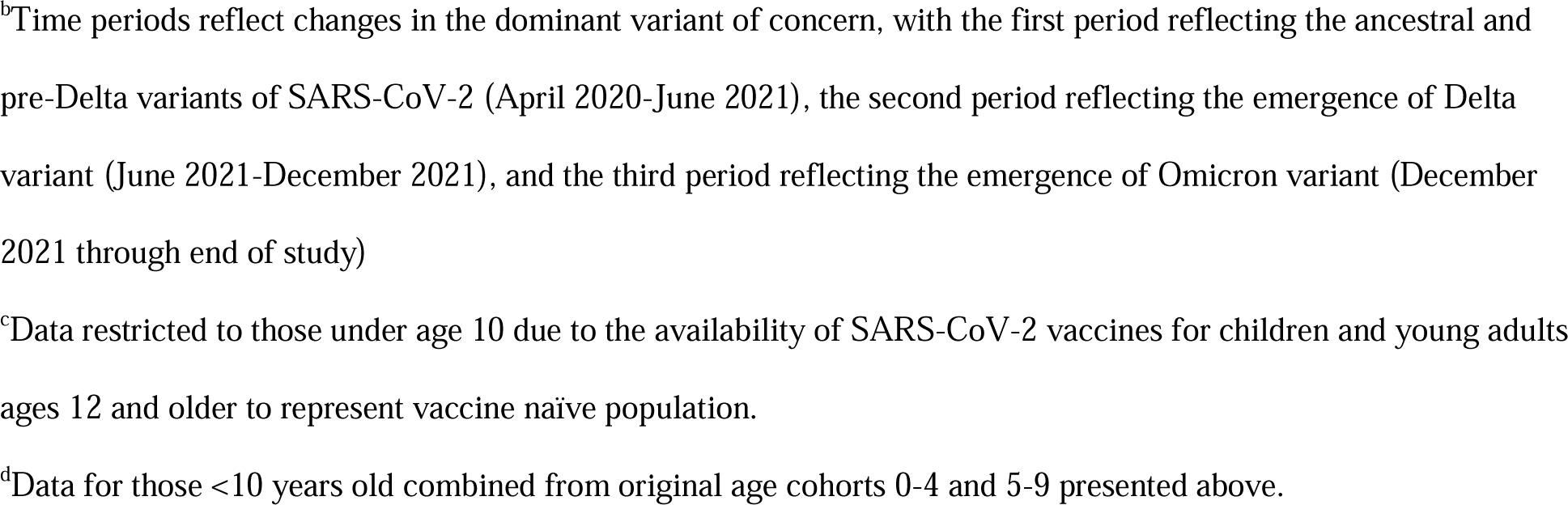
Seroprevalence by timepoint.

| **Phase 1** | **Total N (% of**<br>**time period)** | **N positive** | **Seroprevalence** | **95% CI** |
| --- | --- | --- | --- | --- |
| **November 2020 - May 2021** |  |  |  |  |
| **(Ancestral and pre-Delta**<br>**variants)** |  |  |  |  |
| 0-4 years | 335 (15.9) | 11 | 3.28 | 1.84, 5.78 |
| 5-9 years | 401 (19.0) | 17 | 4.24 | 2.66, 6.68 |
| All 0-9 years | 736 | 28 | 3.80 | 2.65, 5.44 |
| 10-14 years | 455 (21.6) | 14 | 3.08 | 1.84, 5.1 |
| 15-19 years | 458 (21.7) | 18 | 2.50 | 2.50, 6.13 |
| 20-24 years | 461 (21.8) | 33 | 5.14 | 5.14, 9.88 |
| **Phase 2** |  |  |  |  |
| **June 2021 – December 2021** |  |  |  |  |
| **(Delta variant)<sup>c</sup>** |  |  |  |  |
| 0-4 years | 190 (32.3) | 16 | 8.42 | 5.25, 13.24 |
| 5-9 years | 399 (67.7) | 22 | 5.51 | 3.67, 8.21 |
| All 0-9 years<sup>d</sup> | 589 | 38 | 6.45 | 4.74, 8.73 |
| **January 2022 – May 2022** |  |  |  |  |
| **(Omicron variant)<sup>c</sup>** |  |  |  |  |
| 0-4 years | 73 (88.0) | 16 | 21.92 | 13.97, 32.68 |
| 5-9 years | 10 (12.1) | 6 | 60.00 | 31.27, 83.18 |
| All 0-9 years<sup>d</sup> | 83 | 22 | 26.51 | 18.2, 36.89 |

<sup>a</sup>95% CI = 95<sup>th</sup> percentile confidence interval; N = number
<sup>b</sup>Time periods reflect changes in the dominant variant of concern, with the first period reflecting the ancestral and pre-Delta variants of SARS-CoV-2 (April 2020-June 2021), the second period reflecting the emergence of Delta variant (June 2021-December 2021), and the third period reflecting the emergence of Omicron variant (December 2021 through end of study)
<sup>c</sup>Data restricted to those under age 10 due to the availability of SARS-CoV-2 vaccines for children and young adults ages 12 and older to represent vaccine naïve population.
<sup>d</sup>Data for those <10 years old combined from original age cohorts 0-4 and 5-9 presented above.

Young adults aged 20-24 years and infants under age five had the highest overall seropositivity rates of 7.2% (95% CI: 5.2, 9.9) and 7.0% (95% CI: 5.2, 9.3), respectively (p=0.009) (Table 1). There were no significant differences in seropositivity between sexes or between those with or without pre-existing medical conditions (Table 1). South Asian participants had the highest seropositivity (13.5%; 95% CI: 7.5, 23.1) among all ethnic groups (Table 1). Individuals who had travelled internationally had a higher seropositivity rate than those who had travelled within Canada or not travelled (7.7% vs 5.1%; p=0.046) Table 1). There was no significant difference in seropositivity among participants working or attending school compared with those who did not.

In our first multivariable model (Table 3), children aged 10-14 years had lower odds of seropositivity compared with those aged under five years (OR: 0.41; 95% CI: 0.22-0.77). In contrast, South Asian participants (compared to white; OR: 2.95; 95% CI: 1.44, 6.04) and those who had travelled internationally since January 2020 (compared to those with no travel; OR: 1.62; 95% CI: 1.03, 2.55) had higher odds of seropositivity. In our second multivariable model (Table 3) of children aged under ten years, seropositivity rose significantly in later time periods compared to during circulation of the ancestral and pre-Delta variant of concern, with an OR of 1.87 (95% CI: 1.1, 3.1) during the Delta time period and 9.57 (95% CI: 5.1, 17.9) during the Omicron time period.

**Table 3:**
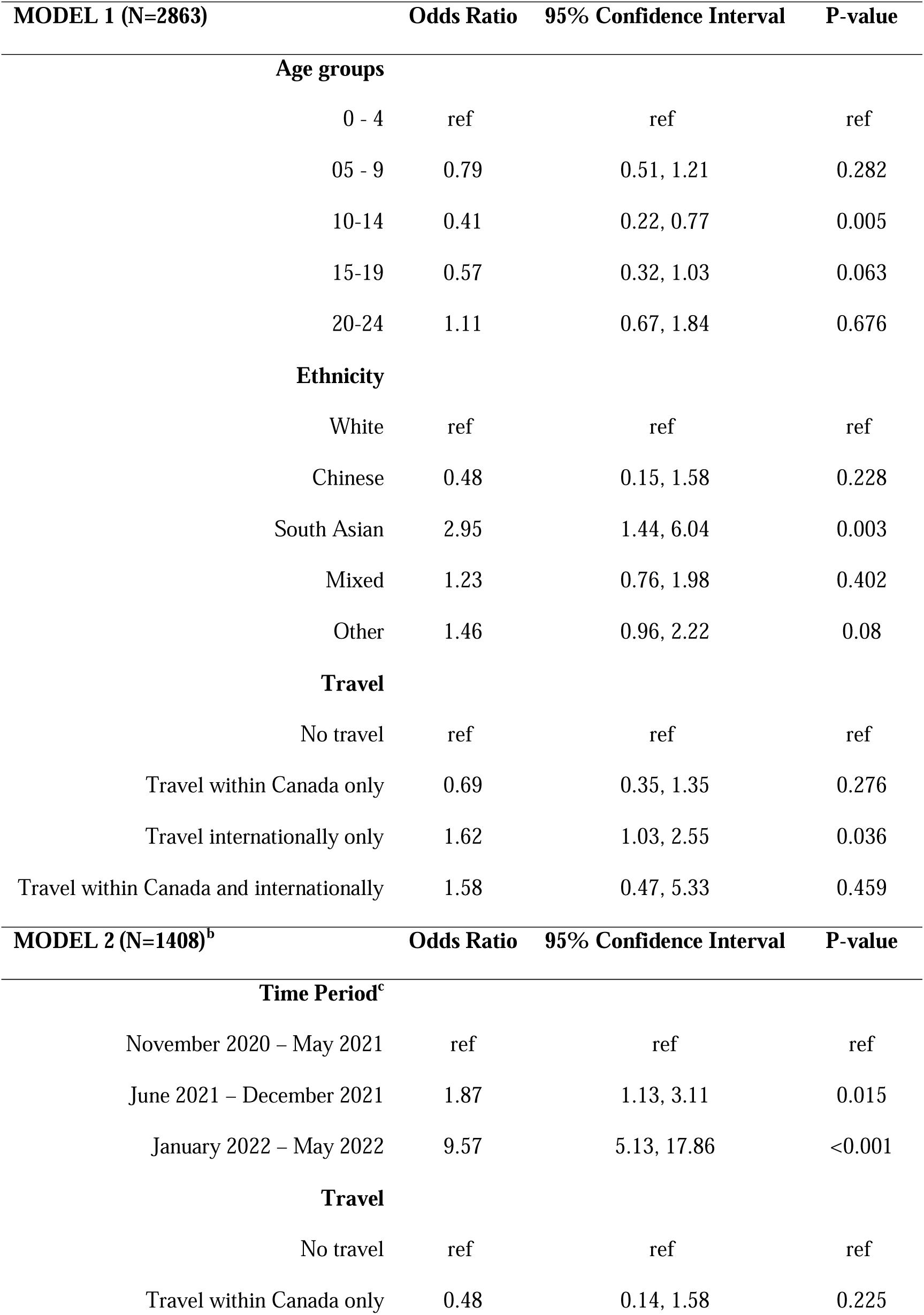
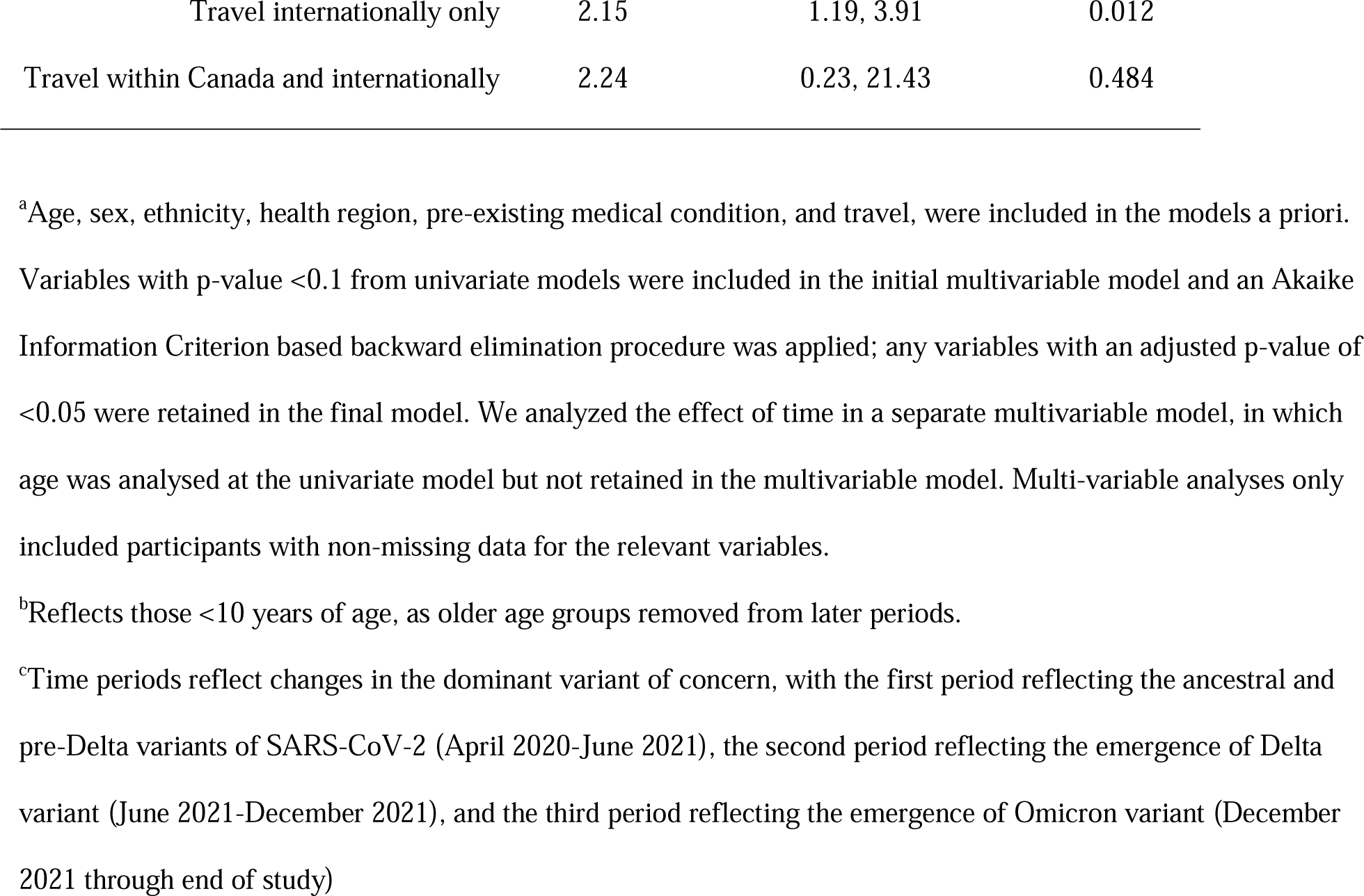
Multivariable analysis.

### Comparison of reported SARS-CoV-2 testing history and seropositivity data

Of the 156 participants who were anti-S IgG positive, 120 did not report SARS-COV-2 positive testing history. Among the 79 who did report prior COVID-19 testing history, 43 participants (54%) reported only negative results (Supplementary table 3); 76 participants had never been tested. Only 2/38 participants (5%) who reported a positive SARS-CoV-2 test in the past were found to be seronegative.

### Anti-S IgG concentrations

Among seropositive participants, the anti-S IgG geometric mean concentration (GMC) was 660 AU/mL (95% CI: 502, 869). Antibody concentrations were higher among those with reported a positive COVID-19 testing history compared with those with a negative testing history. Specifically, of those with positive anti-S IgG, the GMC was 895 AU/mL (95% CI: 615, 1303) for those who reported a positive testing history, compared with 508 AU/mL (95% CI: 341, 758) for those who reported a negative testing history. Of those who reported no history of SARS-CoV-2 test, anti-S IgG GMCs were 434.58 (307.67, 613.85).

## DISCUSSION

This analysis, which evaluated SARS-CoV-2 seropositivity in unvaccinated children through May 2022, demonstrated a low seropositivity rate in 2021, despite returns to in-person schooling. Seropositivity was increased in early 2022, associated with the Omicron variant. These results support the idea that targeted preventive interventions can allow children and youth to safely continue social and educational activities in person, thus minimizing psychosocial impacts. [12–14] Despite evaluation of potential exposures, the main risks of seropositivity reflected demographic factors (i.e., age, ethnicity). We speculate the reported high rates of adherence to public health measures such as masking may have mitigated risk in congregate settings. These results are also in keeping with other seroprevalence and contact tracing studies conducted in BC school settings that showed low rates of SARS-CoV-2 transmission.[15–19]

We demonstrated a higher seropositivity among infants and young adults compared with school-aged children. Our findings contrast with other cohorts in which seropositivity has been reported. A British study completed in 2020 demonstrated no difference among age groups, while a Greek study completed over 2020-2021 found higher rates among 4-6 year-olds particularly earlier in the pandemic and lowest rates among those 0-1 years, whereas we found higher rates in our youngest (0-4 years) and oldest (20-24 years) participants despite school-aged children returning to in-person schooling during our study period.[3, 20] However, our data generally correlated with the overall distribution of SARS-CoV-2 cases in BC since the onset of the pandemic, with rates among 20-29 year-olds much higher than those in ages 10-19 or those under age 10.

Our study demonstrated higher seropositivity among South Asian participants compared with participants with other ethnicities. Neighbourhood-level analysis of COVID-19 cases in Ontario identified several socio-demographic variables associated with higher incidence and mortality, and particularly highlight neighborhoods with high proportions of immigrants, racialized populations, and specifically areas with higher proportions of Black, Latin American, and South Asian residents.[21–24] These elevated rates were directly seen among those in the lowest income quintiles, and related to housing conditions, precarious occupations, and rooted in other systemic inequalities.[21, 24, 25] We found similar trends among South Asian participants independent of age, while we cannot comment on seropositivity rates among Black and LatinX British Columbians as they were inadequately represented.

This study demonstrated the utility of DBS to effectively reach children and young adults through a convenient sampling tool and perform serial seroprevalence testing. We demonstrated many seropositive participants that reported no previous SARS-CoV-2 test. As future novel infections develop, there is a need to identify the disease burden across the pediatric age range, particularly among young children, whom we have demonstrated to have had higher rates of infection, and for whom traditional diagnostics requiring invasive sampling can be challenging. Diagnostics that are sensitive and child-friendly are important for adequately assessing the disease burden to allow for appropriate public health guidelines. Our use of DBS across age groups, allowing for diverse cross-provincial sampling and allow for more inclusive sampling of participants in rural and remote communities, highlights the opportunity for use of DBS as a child-friendly tool.

Seropositivity rates were variable across settings and remained low across Canada through mid-2021. Our study identified a higher SARS-CoV-2 infection burden compared to provincial rates based on SARS-CoV-2 polymerase chain reaction (PCR) data. While the BCCDC reported a PCR positivity rate of 1.7% in children aged ten years and under through May, 2021, we found a seropositivity of 4% in the same age-specific cohort over a similar time period.[26] The BCCDC reported PCR positivity rates of approximately 4% by early January 2022 and 6% by May 2022 in the same age group, while we found a seropositivity of 6.5% and 26.5 in those time periods, respectively.[27, 28] In contrast, a series of cross-sectional serosurveys study in BC reported similar estimates in January 2021 to our data in snapshots between November 2020 through May 2021, but our later seropositivity estimates were substantially lower than their reported infection-induced seroprevalence.[6] However, differences between the two studies likely reflect differences in study design and sampled population, limiting opportunities for direct comparison.

This analysis has numerous strengths. Our study population was largely representative of the provincial multi-ethnic makeup; while South Asians make up 5.8% of the BC population and Chinese make up 7.9%, they made up 2.6% and 3.7% of our cohort respectively, with an additional 14% reporting mixed ethnicity.[29] Our study also contains substantial self-reported data of illnesses, behaviours and symptoms, along with seropositivity data. We were able to sample a large cohort of participants regardless of geographic proximity to a SARS-CoV-2 testing centre. We had low rates of missing data for core demographics, including self-reported ethnicity. Furthermore, the use of DBS had good sensitivity for detecting SARS-CoV-2 infection, with few participants reporting a positive test having a negative serology on DBS. However, this study also has several limitations. Those living in the north and outside of the lower mainland, Indigenous children and young adults, and participants who were not comfortable completing an English-language survey, were under-represented. While we aimed to design the study to be accessible across socioeconomic statuses (SES), participants of a low SES background are often under-represented in research. Other Canadian studies have highlighted that these individuals may have been disproportionately affected by the pandemic. Our recruitment methods may have led to sampling of healthier participants than the general population.[24, 25] As these participants represents a snapshot in time, it does not capture the rate at which seropositivity may wane over time in individuals and does not capture the potentially improved hybrid immunity provided by vaccination and infection. The wide time periods reported here do not capture more subtle changes in seropositivity over the course of the reported time periods. Youth over age ten were not able to be included across both phases, limiting our ability to assess these age groups later in the pandemic. Finally, while our DBS samples identified nearly all participants (94%) who reported a prior positive SARS-CoV-2 testing, it may be less sensitive than alternate diagnostics. However, due to its relative ease of use compared to acquiring serum specimens, population-wide estimates based on DBS sampling may enable representative seroprevalence estimates across large geographic areas; further direct comparison studies would be helpful for evaluating these different approaches.

## CONCLUSIONS

In this observational study, we found an overall seropositivity of 5.5% among children and young adults under the age of 25 in British Columbia between November 2020 and May 2022, with higher seropositivity rates among infants ages 0-4 and young adults ages 20-24. Ongoing seropositivity tracking will help understand the epidemiological changes of SARS-CoV-2 variants. Our results reinforce the need for improved surveillance and reporting mechanisms that account for age-dependent disease to better understand pandemic dynamics. Though vaccination among children and young adults remains important, our data provide additional reassurance that widespread transmission did not occur among susceptible children that continued in-person activities through much of the pandemic.

## Data Availability

All data produced in the present study are available upon reasonable request to the authors

## Abbreviations

Anti-S: anti-spike
BC: British Columbia
BCCDC: BC Centre for Disease Control
CI: confidence intervals
COVID-19: Coronavirus disease 2019
DBS: Dried blood spot
GMC: geometric mean concentration
IgG: immunoglobulin G
MSD: Meso Scale Discovery
PCR: polymerase chain reaction
SARS-CoV-2: severe acute respiratory syndrome coronavirus 2
SES: socioeconomic status
STROBE: Strengthening the reporting of observational studies in epidemiology

