## Supplementary table 2 for "Factors associated with severe acute respiratory syndrome-related coronavirus 2 infection in unvaccinated children and young adults"

**SUPPLEMENTARY TABLE 2: Infection prevention and control behaviours**

| **Question^1^** | **Overall Mean score** | **N less confident** | **N neutral (scale median)** | **N confident** |
| --- | --- | --- | --- | --- |
| Hand washing or use of sanitizer in indoor spaces | 4.46 | 79 | 186 | 2597 |
| Maintain a distance of 2 meters from members of the public in indoor spaces | 4.00 | 68 | 454 | 2341 |
| Wear a mask or face covering in indoor spaces | 4.17 | 305 | 248 | 2308 |

^1^Each question was ranked on a 5-point Likert scale, with 5 representing use every time, and 1 representing never using these behaviours. Responses were divided into three: scores 4 or 5 (most of the time or every time), neutral scores of 3, and scores of 1 or 2 (rarely or never).
