## Supplementary table 3 for "Factors associated with severe acute respiratory syndrome-related coronavirus 2 infection in unvaccinated children and young adults"

**SUPPLEMENTARY TABLE 3: Comparison of acute SARS-CoV-2 testing results and seropositivity**

| **Characteristic** | **Negative COVID-19 test, negative serology N (%)**  **N=1059** | **Negative COVID-19 test, positive serology N (%)**  **N=43** | **Positive COVID-19 test, negative serology N (%)**  **N=2** | **Positive COVID-19 test, positive serology N (%)**  **N=36** |
| --- | --- | --- | --- | --- |
| **Age** |  |  |  |  |
| 0-4 | 256 (24.2) | 12 (27.9) | 1 (50) | 15 (41.7) |
| 5-9 | 398 (37.6) | 12 (27.9) | 1 (50) | 12 (33.3) |
| 10-14 | 138 (13) | 3 (7) | 0 | 1 (2.8) |
| 15-19 | 119 (11.2) | 4 (9.3) | 0 | 4 (11.1) |
| 20-24 | 148 (14) | 12 (27.9) | 0 | 4 (11.1) |
| **Sex** |  |  |  |  |
| Male | 508 (48) | 17 (39.5) | 0 | 18 (50) |
| Female | 551 (52) | 26 (60.5) | 2 (100) | 18 (50) |
| **Gender** |  |  |  |  |
| Man/boy | 506 (47.8) | 17 (39.5) | 0 | 18 (50) |
| Woman/Girl | 540 (51) | 26 (60.5) | 2 (100) | 18 (50) |
| Non-binary, genderqueer, agender, two spirit, other | 13 (1.2) | 0 | 0 | 0 |
| **Ethnicity** |  |  |  |  |
| White | 632 (59.7) | 23 (53.5) | 2 (100) | 21 (58.3) |
| Chinese | 35 (3.3) | 1 (2.3) | 0 | 0 |
| South Asian | 23 (2.2) | 3 (7) | 0 | 1 (2.8) |
| Mixed | 145 (13.7) | 6 (14) | 0 | 4 (11.1) |
| Other | 224 (21.2) | 10 (23.3) | 0 | 10 (27.8) |
| **Health region** |  |  |  |  |
| Vancouver Coastal | 340 (32.1) | 16 (37.2) | 0 | 10 (27.8) |
| Fraser | 287 (27.1) | 14 (32.6) | 1 (50) | 14 (38.9) |
| Interior | 55 (5.2) | 3 (7) | 0 | 1 (2.8) |
| Northern | 20 (1.9) | 0 | 0 | 0 |
| Island | 93 (8.8) | 3 (7) | 1 (50) | 1 (2.8) |
| **Pre-existing medical condition** |  |  |  |  |
| No medical condition | 872 (82.3) | 39 (90.7) | 1 (50) | 30 (83.3) |
| Medical condition | 187 (17.7) | 4 (9.3) | 1 (50) | 6 (16.7) |
| **Participant (child) leaving home** |  |  |  |  |
| Work | 195 (18.4) | 11 (25.6) | 0 | 4 (11.1) |
| School | 623 (58.8) | 25 (58.1) | 1 (50) | 18 (50) |
| Daycare | 210 (19.8) | 6 (14) | 1 (50) | 6 (16.7) |
| None | 24 (2.3) | 1 (2.3) | 0 | 1 (2.8) |
| Other | 193 (18.2) | 4 (9.3) | 0 | 6 (16.7) |
| **Since January 2020, travel outside of BC** |  |  |  |  |
| Travel within Canada | 109 (10.3) | 7 (16.3) | 1 (50) | 1 (2.8) |
| Travel outside of Canada | 157 (14.8) | 9 (20.9) | 0 | 8 (22.2) |
| No travel | 610 (57.6) | 21 (48.8) | 1 (50) | 19 (52.8) |
| **Exposure to contact with a positive COVID-19 test** |  |  |  |  |
| Yes | 196 (18.5) | 23 (53.5) | 1 (50) | 31 (86.1) |
| No | 665 (62.8) | 16 (37.2) | 1 (50) | 2 (5.6) |
| Unknown | 198 (18.7) | 4 (9.3) | 0 | 3 (8.3) |
