## Supplementary figure 1 for "Factors associated with severe acute respiratory syndrome-related coronavirus 2 infection in unvaccinated children and young adults"

**Supplemental Digital Content 1: Timeline of COVID-19 pandemic and provincial milestones through June 2022 by SARS-CoV-2 variant of concern^1^**

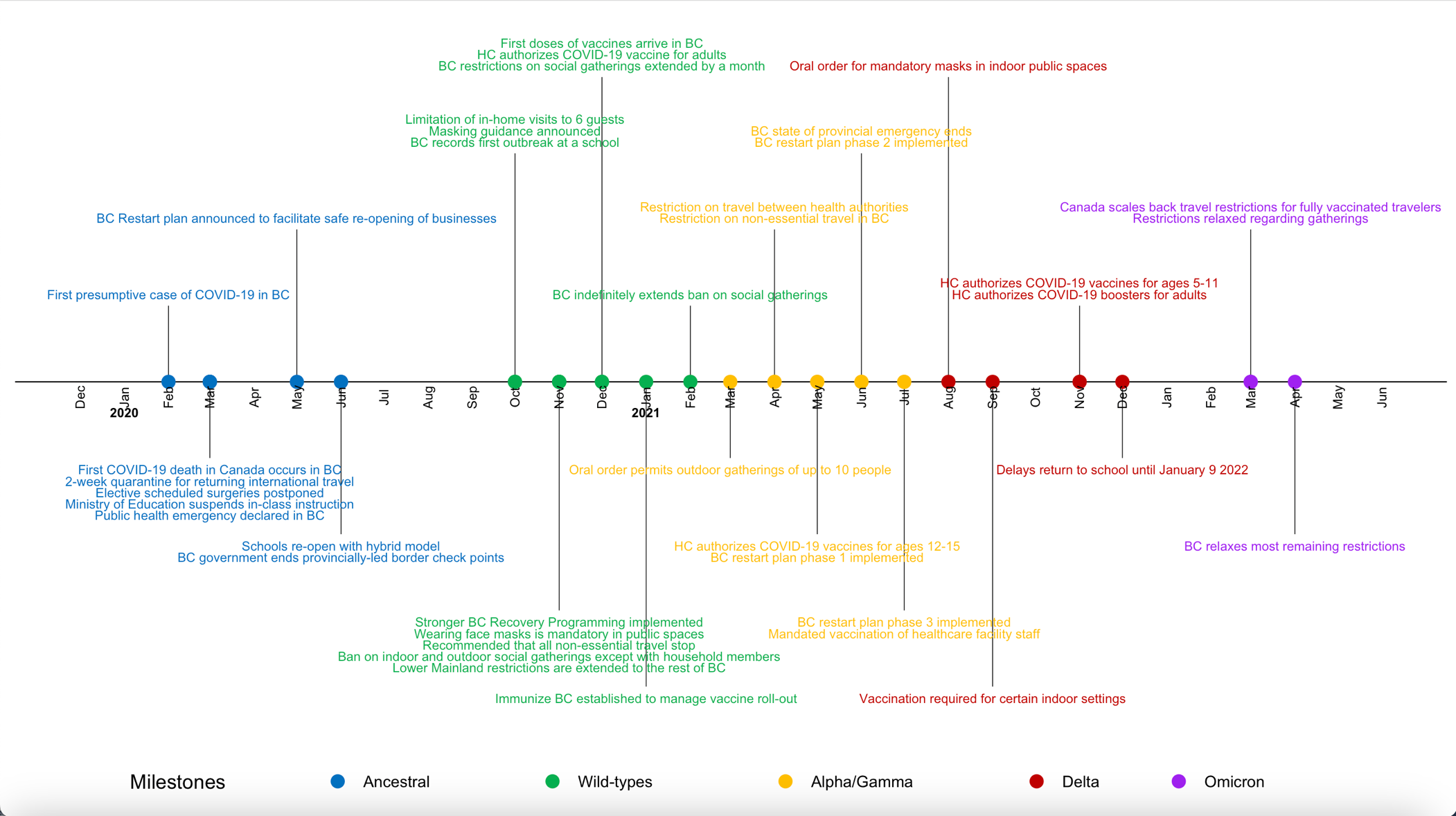

^1^BC refers to British Columbia; HC refers to Health Canada
