## Supplementary figure 2 for "Factors associated with severe acute respiratory syndrome-related coronavirus 2 infection in unvaccinated children and young adults"

#### Baseline Survey

Record ID:

**BACKGROUND INFORMATION**

Please answer the following questions about yourself.

PARENTS: if you are answering this survey on behalf of your child, please answer the questions in relation to your child.

**What is your (your child's) age?** ______ ○ Years ○ Months

You (your child) must be under 25 years old to participate in this study. If you (your child) are 25 years old or older, please contact the SPRING study team.
If you are entering a child's age, please enter in months if under 2 years of age.

**What sex were you (your child) assigned at birth?**
* must provide value

- ○ Male
- ○ Female
- ○ Intersex
- ○ Prefer to self-describe as: ______
- ○ Prefer not to answer

**What best describes your (your child's) gender:**
* must provide value

- ○ Man/boy
- ○ Woman/girl
- ○ Non-binary, GenderQueer, Agender or similar identity
- ○ Two-spirit
- ○ Prefer to self-describe as: ______
- ○ Prefer not to answer

**What are the first three digits of your postal code?** (e.g. V5H) * must provide value ______

**What best describes your (your child's) ethnicity? *(tick all that apply)***
* must provide value

- ☐ Indigenous (First Nations, Metis, Inuk/Inuit)
- ☐ White
- ☐ Chinese
- ☐ South Asian (East Indian, Pakistani, Sri Lankan, etc)
- ☐ Black (African, African-American)
- ☐ Filipino
- ☐ Latin, Central and South American
- ☐ Southeast Asian (Vietnamese, Cambodian, Malaysian, Laotian, etc.)
- ☐ Arabic
- ☐ West Asian (Iranian, Afghan, etc.)
- ☐ Korean
- ☐ Japanese
- ☐ Alaskan Native
- ☐ Native Hawaiian or other Pacific Islander
- ☐ Other, specify: ______
- ☐ Prefer not to answer

**Do you (your child) have any current medical conditions?**
* must provide value

- ○ Yes
- ○ No

reset

**Please list your (your child's) current medical condition(s):** ______

**Are you (your child) currently taking any prescription medication?**
* must provide value

- ○ Yes
- ○ No
- ○ Prefer not to answer

reset

**Please list the name of the prescription medication(s):** ______

#### Baseline Survey

**Since January 2020, did you (your child) leave the home to go to:**
* must provide value

- ☐ Work *(choose all that apply)*
  - ☐ Healthcare worker
  - ☐ Essential worker (e.g. postal, grocery, delivery)
  - ☐ Personal service provider (e.g. hairdresser/barber, esthetician)
  - ☐ Teacher, Principal, School counselor etc.
  - ☐ Other, specify:
- ☐ School *(choose level of schooling)*
  - ○ Preschool
  - ○ Elementary/Primary
  - ○ Secondary/High school
  - ○ Post secondary (college, university or technical institute)
- ☐ Daycare
- ☐ Other, specify: ________
- ☐ None of the above
- ☐ Prefer not to answer

**In the last 6 months, on average, how many times per week do you (your child) use the following transportation?**

| | never | less than 1/week | 1-2 times/week | 3-4 times/week | 5-6 times/week | daily |
|---|---|---|---|---|---|---|
| **Public transportation** * must provide value | ○ | ○ | ○ | ○ | ○ | ○ |
| **Another's private vehicle (such as taxi, car pooling, ride share)** * must provide value | ○ | ○ | ○ | ○ | ○ | ○ |

reset

#### COVID-19 EXPOSURE INFORMATION

**Since January 2020, have you (your child) travelled outside of BC?**
(tick all that apply)
* must provide value

- ☐ Yes, within Canada
- ☐ Yes, outside Canada
- ☐ No
- ☐ Prefer not to answer

**Which province did you (your child) travel to?**
* must provide value

- ☐ Alberta
- ☐ Manitoba
- ☐ New Brunswick
- ☐ Newfoundland and Labrador
- ☐ Northwest Territories
- ☐ Nova Scotia
- ☐ Nunavut
- ☐ Ontario
- ☐ Prince Edward Island
- ☐ Quebec
- ☐ Saskatchewan
- ☐ Yukon
- ☐ Prefer not to answer

**How many total weeks did you (your child) spend outside of BC but within Canada?** ________
* must provide value

**When did you (your child) last return to BC from within Canada?** ________ Today D-M-Y
* must provide value

**What country/countries did you (your child) travel to?** ________
* must provide value

**How many total weeks did you (your child) spend outside of Canada?** ________
* must provide value

**When did you (your child) last return to BC from outside Canada?** ________ Today D-M-Y
* must provide value

#### Baseline Survey

**Have you (your child) ever had any exposure to a person who tested positive for COVID-19?**
* must provide value

- ○ Yes
- ○ No
- ○ I don't know
- ○ Prefer not to answe

reset

**How many people have you (your child) been exposed to?**
* must provide value

______

**Exposure #1**

**Please identify where the exposure occurred: (check all that apply)**

- ☐ Household member
- ☐ Non-household close contact (friend/other family)
- ☐ Health care setting (e.g. Dr. office, physiotherapis/chiropractor office, hospital)
- ☐ School, college, university
- ☐ Personal service provider (e.g. hair salon/barber, esthetician)
- ☐ Workplace
- ☐ Travel, where:
- ☐ Other, specify: ______
- ☐ Prefer not to answer

**When did this exposure occur?**

**Month:** ______ **Year:** ______

**At the time of exposure, do you think the person was still infectious (this is typically within the first 14 days of their illness)?**

- ○ Yes
- ○ No
- ○ I don't know
- ○ Prefer not to answer

**Exposure #2**

**Please identify where the exposure occurred: (check all that apply)**

- ☐ Household member
- ☐ Non-household close contact (friend/other family)
- ☐ Health care setting (e.g. Dr. office, physiotherapis/chiropractor office, hospital)
- ☐ School, college, university
- ☐ Personal service provider (e.g. hair salon/barber, esthetician)
- ☐ Workplace
- ☐ Travel, where:
- ☐ Other, specify: ______
- ☐ Prefer not to answer

**When did this exposure occur?**

**Month:** ______ **Year:** ______

**At the time of exposure, do you think the person was still infectious (this is typically within the first 14 days of their illness)?**

- ○ Yes
- ○ No
- ○ I don't know
- ○ Prefer not to answer

**Exposure #3**

**Please identify where the exposure occurred: (check all that apply)**

- ☐ Household member
- ☐ Non-household close contact (friend/other family)
- ☐ Health care setting (e.g. Dr. office, physiotherapis/chiropractor office, hospital)
- ☐ School, college, university
- ☐ Personal service provider (e.g. hair salon/barber, esthetician)
- ☐ Workplace
- ☐ Travel, where:
- ☐ Other, specify: ______
- ☐ Prefer not to answer

**When did this exposure occur?**

**Month:** ______ **Year:** ______

**At the time of exposure, do you think the person was still infectious (this is typically within the first 14 days of their illness)?**

- ○ Yes
- ○ No
- ○ I don't know
- ○ Prefer not to answer

**Are you (your child) currently in isolation?**

- ○ Yes
- ○ No

reset

**When you (your child) were in indoor public spaces (e.g. shopping, dining) how frequently did you (your child):**

| | Never | Rarely | Some of the time | Most of the time | Every time |
|---|---|---|---|---|---|
| **Wash your hands or use hand sanitizer** * must provide value | ○ | ○ | ○ | ○ | ○ |
| **Maintain a distance of 2 meters from members of the public** * must provide value | ○ | ○ | ○ | ○ | ○ |
| **Wear a mask or other face covering** * must provide value | ○ | ○ | ○ | ○ | ○ |

reset

reset

#### Baseline Survey

##### HOUSEHOLD DETAILS

**Other than yourself (your child), how many people live in your home since January 2020?**
**If you (your child) are a student currently living in student residence, please complete the following table for each roommate you have.**

* must provide value

**Please complete the following table(s) for each household member:**

**Household member #1**

| | |
|---|---|
| **Age in years** (enter as "0" for children less than 1yr old) | |
| **In the last 6 months, did this household member leave the home to go to:** (tick all that apply) | ☐ Work *(choose all jobs in the last 6 months)*<br>☐ Healthcare worker<br>☐ Essential worker (e.g. postal, grocery, delivery)<br>☐ Personal service provider (e.g. hairdresser/barber, esthetician)<br>☐ Teacher, Principal, School counsellor etc.<br>☐ Other, specify:<br>☐ School *(choose level of schooling)*<br>○ Preschool<br>○ Elementary/Primary<br>○ Secondary/High school<br>○ Post secondary (college, university or technical institute)<br>reset<br>☐ Daycare<br>☐ Other, specify:<br>☐ No, none of the above |

**Household member #2**

| | |
|---|---|
| **Age in years** (enter as "0" for children less than 1yr old) | |
| **In the last 6 months, did this household member leave the home to go to:** (tick all that apply) | ☐ Work *(choose all jobs in the last 6 months)*<br>☐ School *(choose level of schooling)*<br>☐ Daycare<br>☐ Other, specify:<br>☐ No, none of the above |

⋮

**Household member #9**

| | |
|---|---|
| **Age in years** (enter as "0" for children less than 1yr old) | |
| **In the last 6 months, did this household member leave the home to go to:** (tick all that apply) | ☐ Work *(choose all jobs in the last 6 months)*<br>☐ School *(choose level of schooling)*<br>☐ Daycare<br>☐ Other, specify:<br>☐ No, none of the above |

#### Baseline Survey

**Since January 2020, have you (your child) been sick?**
* must provide value

- ○ Yes
- ○ No
- ○ Prefer not to answer

reset

**How many times have you (your child) been sick?**
* must provide value

[ ]

| Illness #1 | Illness #2 | Illness #3 |
|---|---|---|
| **When was this illness? (Month/year)** | | |
| [ ▾ ] [ ▾ ] | [ ▾ ] [ ▾ ] | [ ▾ ] [ ▾ ] |
| **Which of the following symptoms did you (your child) experience during this illness? (tick all that apply)** | | |
| ☐ Fever >38 C<br>☐ Chills<br>☐ Cough<br>☐ Shortness of breath<br>☐ Alteration to taste<br>☐ Alteration to smell<br>☐ Nausea<br>☐ Vomiting<br>☐ Diarrhea<br>☐ None of the above<br>☐ Prefer not to answer | ☐ Fever >38?C<br>☐ Chills<br>☐ Cough<br>☐ Shortness of breath<br>☐ Alteration to taste<br>☐ Alteration to smell<br>☐ Nausea<br>☐ Vomiting<br>☐ Diarrhea<br>☐ None of the above<br>☐ Prefer not to answer | ☐ Fever >38?C<br>☐ Chills<br>☐ Cough<br>☐ Shortness of breath<br>☐ Alteration to taste<br>☐ Alteration to smell<br>☐ Nausea<br>☐ Vomiting<br>☐ Diarrhea<br>☐ None of the above<br>☐ Prefer not to answer |
| **How long did this illness last?** | | |
| [ ]<br>○ Days<br>○ Weeks<br>reset | [ ]<br>○ Days<br>○ Weeks<br>reset | [ ]<br>○ Days<br>○ Weeks<br>reset |
| **Did this illness cause you (your child) to miss work or school/daycare?** | | |
| ○ Yes<br>○ No<br>reset | ○ Yes<br>○ No<br>reset | ○ Yes<br>○ No<br>reset |
| **Did you (your child) need to seek medical attention?** | | |
| ○ Yes<br>○ No<br>reset | ○ Yes<br>○ No<br>reset | ○ Yes<br>○ No<br>reset |

**You mentioned above that you sought medical attention, did you (your child) require hospitalization?**
* must provide value

- ○ Yes
- ○ No
- ○ Prefer not to answer

reset

**Did you (your child) require admission to ICU?**
* must provide value

- ○ Yes
- ○ No
- ○ Prefer not to answer

reset

**Are you (your child) still sick?**
* must provide value

- ○ Yes
- ○ No
- ○ Prefer not to answer

reset

**Have you (your child) ever been tested for COVID-19?**
* must provide value

- ○ Yes
- ○ No
- ○ Prefer not to answer

reset

**When were you (your child) last tested?**
* must provide value

mo: [ ▾ ] yr: [ ▾ ]

**What was the result?**
* must provide value

- ○ Positive
- ○ Negative
- ○ Indeterminate/Inconclusive
- ○ I don't know
- ○ Prefer not to answer

reset

#### Baseline Survey

**Have you (your child) been vaccinated against COVID-19?**
* must provide value

- ◯ Yes
- ◯ No

reset

**How many doses of the COVID-19 vaccine have you (your child) received so far?**
* must provide value

- ◯ One dose
- ◯ Two dose
- ◯ More than two dose

**Vaccination date of first dose:**

| Day | Month | Year |
|---|---|---|

* must provide value

**Which vaccine did you (your child) receive?**
* must provide value

- ◯ Pfizer and BioNTech mRNA vaccine
- ◯ Moderna mRNA vaccine
- ◯ AstraZeneca Oxford vaccine
- ◯ Janssen (Johnson & Johnson) vaccine
- ◯ Medicago vaccine
- ◯ Novavax vaccine
- ◯ Sanofi/GSK vaccine
- ◯ Other: ______
- ◯ Don't know

**Vaccination date of second dose:**

| Day | Month | Year |
|---|---|---|

* must provide value

**Which vaccine did you (your child) receive?**
* must provide value

- ◯ Pfizer and BioNTech mRNA vaccine
- ◯ Moderna mRNA vaccine
- ◯ AstraZeneca Oxford vaccine
- ◯ Janssen (Johnson & Johnson) vaccine
- ◯ Medicago vaccine
- ◯ Novavax vaccine
- ◯ Sanofi/GSK vaccine
- ◯ Other: ______
- ◯ Don't know

**Vaccination date of additional dose:**

| Day | Month | Year |
|---|---|---|

* must provide value

**Which vaccine did you (your child) receive?**
* must provide value

- ◯ Pfizer and BioNTech mRNA vaccine
- ◯ Moderna mRNA vaccine
- ◯ AstraZeneca Oxford vaccine
- ◯ Janssen (Johnson & Johnson) vaccine
- ◯ Medicago vaccine
- ◯ Novavax vaccine
- ◯ Sanofi/GSK vaccine
- ◯ Other: ______
- ◯ Don't know

**Next we'd like to ask you some questions about how you feel about vaccines, including a future COVID-19 vaccine:**
**For parents of children < 14 years old, please answer the following questions for yourself:**

| | Very unlikely | Unlikely | Neutral | Somewhat likely | Very likely |
|---|---|---|---|---|---|
| **If a COVID-19 vaccine were to become available to the public, and recommended for you, how likely are you to receive it?** <br>* must provide value | ◯ | ◯ | ◯ | ◯ | ◯ |

#### Baseline Survey

**Do you receive all recommended vaccines when offered to you by health care professionals?**

- Yes
- No
- I don't know

reset

**Thinking back to before the beginning of the pandemic (December 2019), how much did you value vaccines?**

- No value
- Very little value
- Neutral
- A little value
- Valued a lot

reset

**Today, how much do you value vaccines?**

- No value
- Very little value
- Neutral
- A little value
- Valued a lot

reset

**Have you received the influenza vaccine in the past 5 years?**

- Never
- 1-2 times
- 3-4 times
- Every year
- I don't know

reset

*Variable:* kab_next_fluvaccine

**Do you intend to or did you receive the influenza vaccine in the fall of 2020?**

- Yes
- No
- I don't know

reset

@HIDDEN

Add Field | Add Matrix of Fields | Import from Field Bank

*Variable:* kab_last_fluvaccine

**Did you receive the influenza vaccine in the fall of [last year]?**

- Yes
- No
- I don't know

reset

Add Field | Add Matrix of Fields | Import from Field Bank

*Variable:* kab_current_fluvaccine

**Have you received the influenza vaccine in the fall of [current year]?**

- Yes
- No
- I don't know

reset

Add Field | Add Matrix of Fields | Import from Field Bank

*Variable:* kab_intend_fluvaccine *Branching logic:* [kab_current_fluvaccine] = 'N'

**Do you intend to receive the influenza vaccine in [current year]?**

- Yes
- No
- I don't know

reset

#### Baseline Survey

**Please indicate how much you agree or disagree with the following statements:**

| | Strongly disagree | Somewhat disagree | Neutral | Somewhat agree | Strongly agree |
|---|---|---|---|---|---|
| Childhood vaccines are important for a child's health | ○ | ○ | ○ | ○ | ○ |
| Getting vaccines is a good way to protect children from disease | ○ | ○ | ○ | ○ | ○ |
| Childhood vaccines are effective | ○ | ○ | ○ | ○ | ○ |
| Having a child vaccinated is important for the health of others in my community | ○ | ○ | ○ | ○ | ○ |
| All childhood vaccines offered by the BC immunization program in my community are beneficial | ○ | ○ | ○ | ○ | ○ |
| The information I receive about vaccines from the vaccination program is reliable and trustworthy | ○ | ○ | ○ | ○ | ○ |
| Generally, I do what my doctor or health care provider recommends about vaccines | ○ | ○ | ○ | ○ | ○ |
| New vaccines carry more risks than older vaccines | ○ | ○ | ○ | ○ | ○ |
| I am concerned about potential serious adverse effects of vaccines | ○ | ○ | ○ | ○ | ○ |
| Children do not need vaccines for diseases that are not common anymore | ○ | ○ | ○ | ○ | ○ |
| The risk associated with diseases is larger than the risk associated with vaccinations | ○ | ○ | ○ | ○ | ○ |

#### Baseline Survey

**Assuming that a safe and effective COVID-19 vaccine was available and recommended, please indicate how much you agree or disagree with the following statements:**

| | Strongly disagree | Disagree | Neutral | Agree | Strongly agree |
|---|---|---|---|---|---|
| A COVID-19 vaccine would be beneficial<br>* must provide value | ○ | ○ | ○ | ○ | ○ |
| A COVID-19 vaccine would be beneficial for children<br>* must provide value | ○ | ○ | ○ | ○ | ○ |
| A COVID-19 vaccine would be beneficial for individuals 60-years and older<br>* must provide value | ○ | ○ | ○ | ○ | ○ |
| COVID-19 is a serious illness<br>* must provide value | ○ | ○ | ○ | ○ | ○ |
| A COVID-19 vaccine would be beneficial for the health of my community<br>* must provide value | ○ | ○ | ○ | ○ | ○ |
| A COVID-19 vaccine would be safe<br>* must provide value | ○ | ○ | ○ | ○ | ○ |
| A COVID-19 vaccine would be effective in preventing COVID-19<br>* must provide value | ○ | ○ | ○ | ○ | ○ |
| A COVID-19 vaccine should be mandatory<br>* must provide value | ○ | ○ | ○ | ○ | ○ |

**Please answer the following questions, with the assumption that a safe and effective COVID-19 vaccine is available and recommended.**

| | Strongly disagree | Disagree | Neutral | Agree | Strongly agree |
|---|---|---|---|---|---|
| Most people who are important to me would think that I should receive the COVID-19 vaccine | ○ | ○ | ○ | ○ | ○ |
| People who are important to me would expect me to receive the COVID-19 vaccine | ○ | ○ | ○ | ○ | ○ |
| I would feel under social pressure to receive a COVID-19 vaccine | ○ | ○ | ○ | ○ | ○ |
| Everyone I know would get the COVID-19 vaccine | ○ | ○ | ○ | ○ | ○ |
| My family physician's (or other primary Health Care Provider's) opinion is important to me | ○ | ○ | ○ | ○ | ○ |
| What the BC Public Health Officer recommends is important to follow | ○ | ○ | ○ | ○ | ○ |
| What my coworkers think is important to me | ○ | ○ | ○ | ○ | ○ |
| What my employer/work institution thinks is important to me | ○ | ○ | ○ | ○ | ○ |
| What my school/children's school thinks is important to me | ○ | ○ | ○ | ○ | ○ |
| What my friends think is important to me | ○ | ○ | ○ | ○ | ○ |
| What my family thinks is important to me | ○ | ○ | ○ | ○ | ○ |

#### Baseline Survey

Please answer the following questions, with the assumption that a safe and effective COVID-19 vaccine is available and recommended.

| | Strongly disapprove | Disapprove | Neutral | Approve | Strongly approve |
|---|---|---|---|---|---|
| My family physician (or other primary Health Care Provider) would [approve/disapprove] of me receiving a COVID-19 vaccine | ○ | ○ | ○ | ○ | ○ |
| The BC Public Health Officer would [approve/disapprove] of me receiving the COVID-19 vaccine | ○ | ○ | ○ | ○ | ○ |
| My coworkers would [approve/disapprove] of me receiving the COVID-19 vaccine | ○ | ○ | ○ | ○ | ○ |
| My employer/work institution would [approve/disapprove] of me receiving the COVID-19 vaccine | ○ | ○ | ○ | ○ | ○ |
| The educational institution (elementary/high school/college/university) that I or my children attend/are associated with would [approve/disapprove] of me receiving the COVID-19 vaccine | ○ | ○ | ○ | ○ | ○ |
| My friends would [approve/disapprove] of me receiving the COVID-19 vaccine | ○ | ○ | ○ | ○ | ○ |
| My family would [approve/disapprove] of me receiving the COVID-19 vaccine | ○ | ○ | ○ | ○ | ○ |

For the next 3 questions: If a COVID-19 vaccine was publicly funded and available, like the flu shot, ...

| | Strongly disagree | Disagree | Neutral | Agree | Strongly agree |
|---|---|---|---|---|---|
| ...It would be difficult to receive the COVID-19 vaccine | ○ | ○ | ○ | ○ | ○ |
| ... I could easily receive a COVID-19 vaccine if I wanted to | ○ | ○ | ○ | ○ | ○ |
| ... It would be completely up to me whether I received the COVID-19 vaccine | ○ | ○ | ○ | ○ | ○ |

How much control do you feel you would have over whether you receive a COVID-19 vaccine?

- ○ Very little control
- ○ Not much control
- ○ Neutral
- ○ Some control
- ○ A lot of control

Please make sure you click the "submit" button to complete your survey. Your survey responses will not be registered if you do not click "submit".

### Baseline Survey-household

**Record ID:**

**BACKGROUND INFORMATION**

PARENTS: Please answer the questions in relation to your child

**What is your child's age?** [ ] ○ Years ○ Months

Your child must be under 25 years old to participate in this study. If your child is 25 years old or older, please contact the SPRING study team.
If your child is under 2 years of age, please enter the age in months.

**What sex was your child assigned at birth?**
* must provide value

- ○ Male
- ○ Female
- ○ Intersex
- ○ Prefer to self-describe as: [ ]
- ○ Prefer not to answer

**What best describes your child's gender:**
* must provide value

- ○ Man/boy
- ○ Woman/girl
- ○ Non-binary, GenderQueer, Agender or similar identity
- ○ Two-spirit
- ○ Prefer to self-describe as: [ ]
- ○ Prefer not to answer

**What are the first three digits of your child's postal code?**
(e.g. V5H) * must provide value
[ ]

**What best describes your child's ethnicity?**
***(tick all that apply)***
* must provide value

- ☐ Indigenous (First Nations, Metis, Inuk/Inuit)
- ☐ White
- ☐ Chinese
- ☐ South Asian (East Indian, Pakistani, Sri Lankan, etc)
- ☐ Black (African, African-American)
- ☐ Filipino
- ☐ Latin, Central and South American
- ☐ Southeast Asian (Vietnamese, Cambodian, Malaysian, Laotian, etc.)
- ☐ Arabic
- ☐ West Asian (Iranian, Afghan, etc.)
- ☐ Korean
- ☐ Japanese
- ☐ Alaskan Native
- ☐ Native Hawaiian or other Pacific Islander
- ☐ Other, specify: [ ]
- ☐ Prefer not to answer

**Does your child have any current medical conditions?**
* must provide value

- ○ Yes
- ○ No

reset

**Please list your child's current medical condition(s):** [ ]

**Is your child currently taking any prescription medication?**
* must provide value

- ○ Yes
- ○ No
- ○ Prefer not to answer

reset

**Please list the name of the prescription medication(s):** [ ]

#### Baseline Survey-household

**Since January 2020, did your child leave the home to go to:**
* must provide value

- ☐ Work *(choose all that apply)*
  - ☐ Healthcare worker
  - ☐ Essential worker (e.g. postal, grocery, delivery)
  - ☐ Personal service provider (e.g. hairdresser/barber, esthetician)
  - ☐ Teacher, Principal, School counselor etc.
  - ☐ Other, specify:
- ☐ School *(choose level of schooling)*
  - ○ Preschool
  - ○ Elementary/Primary
  - ○ Secondary/High school
  - ○ Post secondary (college, university or technical institute)
- ☐ Daycare
- ☐ Other, specify: ____
- ☐ None of the above
- ☐ Prefer not to answer

**In the last 6 months, on average, how many times per week does your child use the following transportation?**

| | never | less than 1/week | 1-2 times/week | 3-4 times/week | 5-6 times/week | daily | |
|---|---|---|---|---|---|---|---|
| **Public transportation** * must provide value | ○ | ○ | ○ | ○ | ○ | ○ | reset |
| **Another's private vehicle (such as taxi, car pooling, ride share)** * must provide value | ○ | ○ | ○ | ○ | ○ | ○ | reset |

**COVID-19 EXPOSURE INFORMATION**

**Since January 2020, has your child travelled outside of BC?**

(tick all that apply)
* must provide value

- ☐ Yes, within Canada
- ☐ Yes, outside Canada
- ☐ No
- ☐ Prefer not to answer

**Which province did your child travel to?**
* must provide value

- ☐ Alberta
- ☐ Manitoba
- ☐ New Brunswick
- ☐ Newfoundland and Labrador
- ☐ Northwest Territories
- ☐ Nova Scotia
- ☐ Nunavut
- ☐ Ontario
- ☐ Prince Edward Island
- ☐ Quebec
- ☐ Saskatchewan
- ☐ Yukon
- ☐ Prefer not to answer

**How many total weeks did your child spend outside of BC but within Canada?** ____
* must provide value

**When did your child last return to BC from within Canada?** ____ Today D-M-Y
* must provide value

**What country/countries did your child travel to?** ____
* must provide value

**How many total weeks did your child spend outside of Canada?** ____
* must provide value

**When did your child last return to BC from outside Canada?** ____ Today D-M-Y
* must provide value

#### Baseline Survey-household

**Has your child ever had any exposure to a person who tested positive for COVID-19?**
* must provide value

- ○ Yes
- ○ No
- ○ I don't know
- ○ Prefer not to answe

reset

**How many people has your child been exposed to?**
* must provide value

[ ]

##### Exposure #1

**Please identify where the exposure occurred: (check all that apply)**
- ☐ Household member
- ☐ Non-household close contact (friend/other family)
- ☐ Health care setting (e.g. Dr. office, physiotherapis/chiropractor office, hospital)
- ☐ School, college, university
- ☐ Personal service provider (e.g. hair salon/barber, esthetician)
- ☐ Workplace
- ☐ Travel, where:
- ☐ Other, specify: [ ]
- ☐ Prefer not to answer

**When did this exposure occur?**
**Month:** [ ] **Year:** [ ]

**At the time of exposure, do you think the person was still infectious (this is typically within the first 14 days of their illness)?**
- ○ Yes
- ○ No
- ○ I don't know
- ○ Prefer not to answer

##### Exposure #2

**Please identify where the exposure occurred: (check all that apply)**
- ☐ Household member
- ☐ Non-household close contact (friend/other family)
- ☐ Health care setting (e.g. Dr. office, physiotherapis/chiropractor office, hospital)
- ☐ School, college, university
- ☐ Personal service provider (e.g. hair salon/barber, esthetician)
- ☐ Workplace
- ☐ Travel, where:
- ☐ Other, specify: [ ]
- ☐ Prefer not to answer

**When did this exposure occur?**
**Month:** [ ] **Year:** [ ]

**At the time of exposure, do you think the person was still infectious (this is typically within the first 14 days of their illness)?**
- ○ Yes
- ○ No
- ○ I don't know
- ○ Prefer not to answer

##### Exposure #3

**Please identify where the exposure occurred: (check all that apply)**
- ☐ Household member
- ☐ Non-household close contact (friend/other family)
- ☐ Health care setting (e.g. Dr. office, physiotherapis/chiropractor office, hospital)
- ☐ School, college, university
- ☐ Personal service provider (e.g. hair salon/barber, esthetician)
- ☐ Workplace
- ☐ Travel, where:
- ☐ Other, specify: [ ]
- ☐ Prefer not to answer

**When did this exposure occur?**
**Month:** [ ] **Year:** [ ]

**At the time of exposure, do you think the person was still infectious (this is typically within the first 14 days of their illness)?**
- ○ Yes
- ○ No
- ○ I don't know
- ○ Prefer not to answer

**Is your child currently in isolation?**
- ○ Yes
- ○ No

reset

**When your child was in indoor public spaces (e.g. shopping, dining) how frequently did your child:**

| | Never | Rarely | Some of the time | Most of the time | Every time |
|---|---|---|---|---|---|
| **Wash hands or use hand sanitizer** * must provide value | ○ | ○ | ○ | ○ | ○ |
| **Maintain a distance of 2 meters from members of the public** * must provide value | ○ | ○ | ○ | ○ | ○ |
| **Wear a mask or other face covering** * must provide value | ○ | ○ | ○ | ○ | ○ |

reset

reset

#### Baseline Survey-household

##### HOUSEHOLD DETAILS

**Other than your child, how many people live in your home since January 2020?**
* must provide value

**Please complete the following table(s) for each household member:**

**Household member #1**

| | |
|---|---|
| **Age in years** (enter as "0" for children less than 1yr old) | |
| **In the last 6 months, did this household member leave the home to go to:** (tick all that apply) | ☐ Work *(choose all jobs in the last 6 months)*<br>☐ Healthcare worker<br>☐ Essential worker (e.g. postal, grocery, delivery)<br>☐ Personal service provider (e.g. hairdresser/barber, esthetician)<br>☐ Teacher, Principal, School counsellor etc.<br>☐ Other, specify:<br>☐ School *(choose level of schooling)*<br>○ Preschool<br>○ Elementary/Primary<br>○ Secondary/High school<br>○ Post secondary (college, university or technical institute)<br>reset<br>☐ Daycare<br>☐ Other, specify:<br>☐ No, none of the above |

**Household member #2**

| | |
|---|---|
| **Age in years** (enter as "0" for children less than 1yr old) | |
| **In the last 6 months, did this household member leave the home to go to:** (tick all that apply) | ☐ Work *(choose all jobs in the last 6 months)*<br>☐ School *(choose level of schooling)*<br>☐ Daycare<br>☐ Other, specify:<br>☐ No, none of the above |

⋮

**Household member #9**

| | |
|---|---|
| **Age in years** (enter as "0" for children less than 1yr old) | |
| **In the last 6 months, did this household member leave the home to go to:** (tick all that apply) | ☐ Work *(choose all jobs in the last 6 months)*<br>☐ School *(choose level of schooling)*<br>☐ Daycare<br>☐ Other, specify:<br>☐ No, none of the above |

#### Baseline Survey-household

**Since January 2020, has your child been sick?**
* must provide value

- ○ Yes
- ○ No
- ○ Prefer not to answer

reset

**How many times has your child been sick?**
* must provide value

[ ]

| Illness #1 | Illness #2 | Illness #3 |
|---|---|---|
| **When was this illness? (Month/year)** | | |
| [ ] [ ] | [ ] [ ] | [ ] [ ] |
| **Which of the following symptoms did your child experience during this illness? (tick all that apply)** | | |
| ☐ Fever >38 C<br>☐ Chills<br>☐ Cough<br>☐ Shortness of breath<br>☐ Alteration to taste<br>☐ Alteration to smell<br>☐ Nausea<br>☐ Vomiting<br>☐ Diarrhea<br>☐ None of the above<br>☐ Prefer not to answer | ☐ Fever >38?C<br>☐ Chills<br>☐ Cough<br>☐ Shortness of breath<br>☐ Alteration to taste<br>☐ Alteration to smell<br>☐ Nausea<br>☐ Vomiting<br>☐ Diarrhea<br>☐ None of the above<br>☐ Prefer not to answer | ☐ Fever >38?C<br>☐ Chills<br>☐ Cough<br>☐ Shortness of breath<br>☐ Alteration to taste<br>☐ Alteration to smell<br>☐ Nausea<br>☐ Vomiting<br>☐ Diarrhea<br>☐ None of the above<br>☐ Prefer not to answer |
| **How long did this illness last?** | | |
| [ ]<br>○ Days<br>○ Weeks<br>reset | [ ]<br>○ Days<br>○ Weeks<br>reset | [ ]<br>○ Days<br>○ Weeks<br>reset |
| **Did this illness cause your child to miss work or school/daycare?** | | |
| ○ Yes<br>○ No<br>reset | ○ Yes<br>○ No<br>reset | ○ Yes<br>○ No<br>reset |
| **Did your child need to seek medical attention?** | | |
| ○ Yes<br>○ No<br>reset | ○ Yes<br>○ No<br>reset | ○ Yes<br>○ No<br>reset |

**You mentioned above that your child sought medical attention, did your child require hospitalization?**
* must provide value

- ○ Yes
- ○ No
- ○ Prefer not to answer

reset

**Did your child require admission to ICU?**
* must provide value

- ○ Yes
- ○ No
- ○ Prefer not to answer

reset

**Is your child still sick?**
* must provide value

- ○ Yes
- ○ No
- ○ Prefer not to answer

reset

**Has your child ever been tested for COVID-19?**
* must provide value

- ○ Yes
- ○ No
- ○ Prefer not to answer

reset

**When was your child last tested?**
* must provide value

mo: [ ] yr: [ ]

**What was the result?**
* must provide value

- ○ Positive
- ○ Negative
- ○ Indeterminate/Inconclusive
- ○ I don't know
- ○ Prefer not to answer

reset

#### Baseline Survey-household

**Has your child been vaccinated against COVID-19?**
* must provide value

- ◯ Yes
- ◯ No

reset

**How many doses of the COVID-19 vaccine has your child received so far?**
* must provide value

- ◯ One dose
- ◯ Two dose
- ◯ More than two dose

**Vaccination date of first dose:**

| Day | Month | Year |
|---|---|---|

* must provide value

**Which vaccine did your child receive?**
* must provide value

- ◯ Pfizer and BioNTech mRNA vaccine
- ◯ Moderna mRNA vaccine
- ◯ AstraZeneca Oxford vaccine
- ◯ Janssen (Johnson & Johnson) vaccine
- ◯ Medicago vaccine
- ◯ Novavax vaccine
- ◯ Sanofi/GSK vaccine
- ◯ Other: ______
- ◯ Don't know

**Vaccination date of second dose:**

| Day | Month | Year |
|---|---|---|

* must provide value

**Which vaccine did your child receive?**
* must provide value

- ◯ Pfizer and BioNTech mRNA vaccine
- ◯ Moderna mRNA vaccine
- ◯ AstraZeneca Oxford vaccine
- ◯ Janssen (Johnson & Johnson) vaccine
- ◯ Medicago vaccine
- ◯ Novavax vaccine
- ◯ Sanofi/GSK vaccine
- ◯ Other: ______
- ◯ Don't know

reset

**Vaccination date of additional dose:**

| Day | Month | Year |
|---|---|---|

* must provide value

**Which vaccine did your child receive?**
* must provide value

- ◯ Pfizer and BioNTech mRNA vaccine
- ◯ Moderna mRNA vaccine
- ◯ AstraZeneca Oxford vaccine
- ◯ Janssen (Johnson & Johnson) vaccine
- ◯ Medicago vaccine
- ◯ Novavax vaccine
- ◯ Sanofi/GSK vaccine
- ◯ Other: ______
- ◯ Don't know

**Please make sure you click the "submit" button to complete your survey. Your survey responses will not be registered if you do not click "submit".**

#### Follow-up Survey

Data Access Group: **[No Assignment]** ?

Invitation status: Survey options

Editing existing Record ID: 3.

Event: **Visit 3 (Arm 1: Arm 1)**

**Record ID:** 3

**BACKGROUND INFORMATION Please answer the following questions about yourself.**

**PARENTS: if you are answering this survey on behalf of your child, please answer the questions in relation to your child.**

**On your last survey completed on _____ 04/28/2022 you indicated that you (your child) had the following medical conditions:**

_____

**Has there been any new or changes to your (your child's) medical conditions?**
* must provide value

- (●) Yes
- ( ) No

reset

**Please list your (your child's) new or changed medical condition(s):** [ ]

**On your last survey completed on _____ 04/28/2022, you indicated that you (your child) were taking following prescription medications:**

_____

**Has there been any new or changes to your (your child's) prescription medications?**
* must provide value

- (●) Yes
- ( ) No
- ( ) Prefer not to answer

reset

**Please list the name of the prescription medication(s):** [ ]

**On your last survey completed on _____ 04/28/2022 you indicated ("checked") that you (your child) left home for the following reasons:**

**Work:** Unchecked - _____

**School:** Unchecked - _____

**Daycare:** Unchecked

**Other:** Unchecked - _____

**None:** Unchecked

**Decline:** Unchecked

#### Follow-up survey continued

**Has your (your child's) job or type of schooling changed?**
* must provide value

- (●) Yes
- ( ) No
- ( ) Prefer not to answer

reset

**Did you (your child) leave the home to go to:**
* must provide value

- [x] Work *(choose all that apply)*
  - [ ] Healthcare worker
  - [ ] Essential worker (e.g. postal, grocery, delivery)
  - [ ] Personal service provider (e.g. hairdresser/barber, esthetician)
  - [ ] Teacher, Principal, School counselor etc.
  - [ ] Other, specify:
  - [ ] Prefer not to answer
- [x] School *(choose level of schooling)*
  - ( ) Preschool
  - ( ) Elementary/Primary
  - ( ) Secondary/High school
  - ( ) Post secondary (college, university or technical institute)
  - ( ) Prefer not to answer

  reset
- [ ] Daycare
- [ ] Other, specify: ____
  50 characters remaining
- [ ] None of the above
- [ ] Prefer not to answer

**Since your last survey completed on _____ 04/28/2022:**

**On average, how many times per week do you (your child) use the following transportation?**

| | never | less than 1/week | 1-2 times/week | 3-4 times/week | 5-6 times/week | daily | |
|---|---|---|---|---|---|---|---|
| **Public transportation** * must provide value | ( ) | ( ) | ( ) | ( ) | ( ) | ( ) | reset |
| **Another's private vehicle (such as taxi, car pooling, ride share)** * must provide value | ( ) | ( ) | ( ) | ( ) | ( ) | ( ) | reset |

**Since your last survey completed on _____ 04/28/2022:**

**Have you (your child) had any exposure to a person who tested positive for COVID-19?**
* must provide value

- (●) Yes
- ( ) No
- ( ) I don't know
- ( ) Prefer not to answer

reset

**How many people have you (your child) been exposed to?**
* must provide value

- ( ) Less than 5 people
- ( ) 5-10 people
- ( ) 10-20 people
- ( ) Greater than 20 people

reset

#### Follow-up survey continued

**Please identify where the most recent exposure occurred: (check all that apply)**

- ☐ Household member
- ☐ Non-household close contact (friend/other family)
- ☐ Health care setting (e.g. Dr. office, physiotherapis/chiropractor office, hospital)
- ☐ School, college, university
- ☐ Daycare
- ☐ Personal service provider (e.g. hair salon/barber, esthetician)
- ☐ Workplace
- ☐ Travel, where:
- ☐ Other, specify:
- ☐ Prefer not to answer

**When did this exposure occur?**

**Month:**

**Year:**

**At the time of exposure, do you think the person was still infectious (this is typically within the first 14 days of their illness)?**

- ○ Yes
- ○ No
- ○ I don't know
- ○ Prefer not to answer

reset

##### HOUSEHOLD DETAILS

**Other than yourself (your child), how many people currently live in your home?**

If you are a parent filling this out for your child, please include yourself in the following table as a household member. If you are a student currently living in student residence, please complete the following table for each roommate you have."

* must provide value

2

**Household member #1**

| |
|---|
| **Age in years** (enter as "0" for children less than 1yr old) |

**Household member #2**

| |
|---|
| **Age in years** (enter as "0" for children less than 1yr old) |

#### Follow-up survey continued

Since your last survey completed on _____ 04/28/2022:

**Have you (your child) been sick that included one or more of the following symptoms:**

- Fever >38°C
- Chills
- Cough
- Shortness of breath
- Alteration to taste
- Alteration to smell
- Nausea
- Vomiting
- Diarrhea

* must provide value

◉ Yes
○ No
○ Prefer not to answer
reset

**How many times have you (your child) been sick?** 4

* must provide value

**Please record details on the three illnesses of most concern**

| Illness #1 | Illness #2 | Illness #3 |
|---|---|---|
| **When was this illness? (Month/year)** | | |
| mo: [ ] yr: [ ] | mo: [ ] yr: [ ] | mo: [ ] yr: [ ] |
| **Did this illness cause you (your child) to miss work or school/daycare?** | | |
| ○ Yes ○ No ○ Prefer not to answer reset | ○ Yes ○ No ○ Prefer not to answer reset | ○ Yes ○ No ○ Prefer not to answer reset |
| **Did you (your child) need to seek medical attention?** | | |
| ○ Yes ○ No ○ Prefer not to answer reset | ○ Yes ○ No ○ Prefer not to answer reset | ○ Yes ○ No ○ Prefer not to answer reset |

#### Follow-up survey continued

**Have you (your child) ever tested positive for COVID-19?**
* must provide value

- (selected) Yes
- No
- Prefer not to answer

reset

**How many times have you (your child) tested positive for COVID-19?**
* must provide value

10

**Please enter your (your child's) POSITIVE tests:**

| | Month of Test | Year of Test | Test Type |
|---|---|---|---|
| Positive COVID-19 Test #1: | | | |
| Positive COVID-19 Test #2: | | | |
| Positive COVID-19 Test #3: | | | |
| Positive COVID-19 Test #4: | | | |
| Positive COVID-19 Test #5: | | | |
| Positive COVID-19 Test #6: | | | |
| Positive COVID-19 Test #7: | | | |
| Positive COVID-19 Test #8: | | | |
| Positive COVID-19 Test #9: | | | |
| Positive COVID-19 Test #10: | | | |

#### Follow-up survey continued

**COVID-19 Vaccinations**

**On previous surveys, you indicated that you (your child) had received the following Dose 1 of the COVID-19 vaccines:**

07-04-2022, Pfizer and BioNTech mRNA vaccine

**Is this dose 1 information correct?**

If the field is empty, and you have recived a dose 1 vaccine, please select "No"

* must provide value

○ Yes
◉ No

reset

| | Vaccination Date | Type of Vaccination |
|---|---|---|
| **Please enter new or updated Dose 1 information:** | [ ] D-M-Y | ○ Pfizer and BioNTech mRNA vaccine<br>○ Moderna mRNA vaccine<br>○ AstraZeneca Oxford vaccine<br>○ Janssen (Johnson & Johnson) vaccine<br>○ Medicago vaccine<br>○ Novavax vaccine<br>○ Sanofi/GSK vaccine<br>○ Other:<br>○ Don't know<br>reset |

**On previous surveys, you indicated that you (your child) had received the following Dose 2 of the COVID-19 vaccines:**

_____, _____

**Is this dose 2 information correct?**

If the field is empty, and you have recived a dose 2 vaccine, please select "No"

* must provide value

○ Yes
◉ No

reset

| | Vaccination Date | Type of Vaccination |
|---|---|---|
| **Please enter new or updated Dose 2 information:** | [ ] D-M-Y | ○ Pfizer and BioNTech mRNA vaccine<br>○ Moderna mRNA vaccine<br>○ AstraZeneca Oxford vaccine<br>○ Janssen (Johnson & Johnson) vaccine<br>○ Medicago vaccine<br>○ Novavax vaccine<br>○ Sanofi/GSK vaccine<br>○ Other:<br>○ Don't know<br>reset |

**On previous surveys, you indicated that you (your child) had received the following Dose 3 of the COVID-19 vaccines:**

_____, _____

**Is this dose 3 information correct?**

If the field is empty, and you have recived a dose 3 vaccine, please select "No"

* must provide value

○ Yes
◉ No

reset

| Please enter new or updated Dose 3 information: | Vaccination Date | Type of Vaccination |
|---|---|---|
| | D-M-Y | ○ Pfizer and BioNTech mRNA vaccine<br>○ Moderna mRNA vaccine<br>○ AstraZeneca Oxford vaccine<br>○ Janssen (Johnson & Johnson) vaccine<br>○ Medicago vaccine<br>○ Novavax vaccine<br>○ Sanofi/GSK vaccine<br>○ Other:<br>○ Don't know<br>reset |

**On previous surveys, you indicated that you (your child) had received the following Dose 4 of the COVID-19 vaccines:**

_____, _____

**Is this dose 4 information correct?**

If the field is empty, and you have recived a dose 4 vaccine, please select "No"

* must provide value

○ Yes
◉ No

reset

| Please enter new or updated Dose 4 information: | Vaccination Date | Type of Vaccination |
|---|---|---|
| | D-M-Y | ○ Pfizer and BioNTech mRNA vaccine<br>○ Moderna mRNA vaccine<br>○ AstraZeneca Oxford vaccine<br>○ Janssen (Johnson & Johnson) vaccine<br>○ Medicago vaccine<br>○ Novavax vaccine<br>○ Sanofi/GSK vaccine<br>○ Other:<br>○ Don't know<br>reset |

**On previous surveys, you indicated that you (your child) had received the following Dose 5 of the COVID-19 vaccines:**

_____, _____

**Is this dose 5 information correct?**

If the field is empty, and you have recived a dose 5 vaccine, please select "No"

* must provide value

○ Yes
◉ No

reset

| Please enter new or updated Dose 5 information: | Vaccination Date | Type of Vaccination |
|---|---|---|
| | D-M-Y | ○ Pfizer and BioNTech mRNA vaccine<br>○ Moderna mRNA vaccine<br>○ AstraZeneca Oxford vaccine<br>○ Janssen (Johnson & Johnson) vaccine<br>○ Medicago vaccine<br>○ Novavax vaccine<br>○ Sanofi/GSK vaccine<br>○ Other:<br>○ Don't know<br>reset |
