## Supplementary figures and images for "Factors associated with severe acute respiratory syndrome-related coronavirus 2 infection in unvaccinated children and young adults"

### Supplementary figure 3

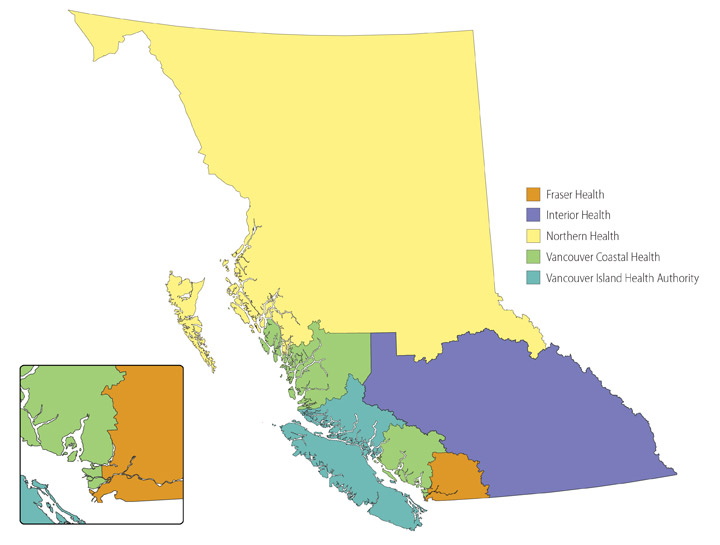
